# CitySEIRCast: An Agent-Based City Digital Twin for Pandemic Analysis and Simulation

**DOI:** 10.1101/2023.12.22.23300481

**Authors:** Shakir Bilal, Wajdi Zaatour, Yilian Alonso Otano, Arindam Saha, Kenneth Newcomb, Kim Soo, Jun Kim, Raveena Ginjala, Derek Groen, Edwin Michael

## Abstract

The COVID-19 pandemic has dramatically highlighted the importance of developing simulation systems for quickly characterizing and providing spatio-temporal forecasts of infection spread dynamics that take specific accounts of the population and spatial heterogeneities that govern pathogen transmission in real-world communities. Developing such computational systems must also overcome the cold-start problem related to the inevitable scarce early data and extant knowledge regarding a novel pathogen’s transmissibility and virulence, while addressing changing population behavior and policy options as a pandemic evolves. Here, we describe how we have coupled advances in the construction of digital or virtual models of real-world cities with an agile, modular, agent-based model of viral transmission and data from navigation and social media interactions, to overcome these challenges in order to provide a new simulation tool, CitySEIRCast, that can model viral spread at the sub-national level. Our data pipelines and workflows are designed purposefully to be flexible and scalable so that we can implement the system on hybrid cloud/cluster systems and be agile enough to address different population settings and indeed, diseases. Our simulation results demonstrate that CitySEIRCast can provide the timely high resolution spatio-temporal epidemic predictions required for supporting situational awareness of the state of a pandemic as well as for facilitating assessments of vulnerable sub-populations and locations and evaluations of the impacts of implemented interventions, inclusive of the effects of population behavioral response to fluctuations in case incidence. This work arose in response to requests from county agencies to support their work on COVID-19 monitoring, risk assessment, and planning, and using the described workflows, we were able to provide uninterrupted bi-weekly simulations to guide their efforts for over a year from late 2021 to 2023. We discuss future work that can significantly improve the scalability and real-time application of this digital city-based epidemic modelling system, such that validated predictions and forecasts of the paths that may followed by a contagion both over time and space can be used to anticipate the spread dynamics, risky groups and regions, and options for responding effectively to a complex epidemic.

## 1 INTRODUCTION

The ongoing COVID-19 pandemic has dramatically highlighted the potential that communicable diseases continue to possess for producing highly destructive global public health and socio-economic threats [7, 31]. The recent epidemics associated with influenza (H1N1) in 2009, the 2011 *Escherichia coli* outbreak in Germany, Ebola in West Africa in 2014, Zika in the Americas in 2016, West Nile virus outbreaks in Europe in 2019, dengue in South America in 2019, and the COVID-19 pandemic that began in 2019 additionally attest to the possibility that new infectious disease outbreaks can emerge at any time anywhere in the world [31, 71]. This threat calls for improved understanding of the invasion and transmission dynamics of epidemic diseases especially in a globalized, interconnected, world, on the one hand, and, on the other, the need for improving predictions of the spread of novel pathogens that also take explicit account of the localized characteristics of settings in order to support effective policy making [15]. They show how making such predictions require addressing the combined effects of deep uncertainty, the impact of intrinsic biology, transmissibility and mutability of a pathogen, the role and outcomes of social heterogeneities and human behavior, and the effects of spatial scale and variability in disease propagation [15, 47, 51, 72].

These contagions, including in the case of COVID-19, have directed attention on the modelling paradigms that are best able to capture the effects of these diverse factors reliably [4, 15, 33, 66, 69]. They have also concentrated focus on the computational workflows and data pipelines required to assemble input data, learn and run models, and provide information to policy makers at the lead times required for making decisions at various spatial scales, including for targeting responses to different subpopulations and risky locations [2, 3, 49, 60, 62]. At its core, these challenges from a disease modelling perspective relate to both the tasks of how best to design dynamic in-silico models whose simulated behavior captures the heterogenous transmission and controllability of novel and extant infectious agents, and the corresponding construction of computational and data systems agile enough to respond to rapidly changing knowledge and policy objectives as these pathogens establish and spread in diverse populations. We indicate that both these challenges present a major barrier to creating and using models for predicting the transmission dynamics of epidemic diseases, especially when extant knowledge of the risk factors and pathogen transmissibility and virulence is limited during the early stage of the invasion of a novel outbreak [50].

The COVID-19 pandemic has also exacerbated and exposed societal inequities for promoting the spread of the contagion as well as for producing variable health and economic outcomes among different subpopulations and geographies [35]. This calls for developing models that are structurally sufficiently detailed for enabling reliable simulations of the effects and outcomes of these societal heterogeneities. Indeed, understanding how interactions between these heterogeneous components of society may operate and affect pathogen transmission in a population will be key to the predictability and controllability of an infectious disease contagion [15, 47, 49]. It will also be critical to assessing and managing societal resilience to large-scale epidemic outbreaks [44]. While several complex data-driven agent-based models (ABMs) have been developed to address aspects of the above modelling challenges [30, 34, 43, 49], three key features remain that impede the development of the systemic modeling approaches required to address epidemic transmission in complex social systems [15]. The first two of these concerns data for informing model development and calibration [37]. These relate firstly to the cold start problem whereby when a new pathogen starts to spread, health departments always need a long lead time to reliably collect sufficient data both on risky subpopulations, activities, and settings as well as on pathogen characteristics, to generate reliable public responses [15, 37, 50]. The second data-related issue is connected with the invariable privacy and confidentiality problems connected with health and population data resulting in both restricted and delayed availability of key spatio-demographic, transmission, and disease-related information required to construct and parameterize the appropriate epidemic models [37]. Finally, the third factor impeding the discovery and construction of epidemiological models for new pathogens is that while there exists a growing list of infectious disease models contributed by independent research activities, these are often not developed for the purpose of reuse or with interoperability in mind meaning that the potential and rapid use of these models or even construction of new models based on existing modelling frameworks is presently severely limited [37].

In recent years, two developments in simulating reality have emerged that may provide a means to overcome the challenges described above for simulating the transmission of infectious pathogens in the real world. The first is the use of a digital twin (DT) as a simulation process for generating a virtual representation of the city’s or community’s physical assets, population characteristics, processes and flows that are connected to all the data related to them and their surrounding environment [16, 17, 52]. As it aims to reflect the whole life cycle process of the corresponding real-world city by tight coupling of the physical and virtual entities and the connections between them [26], such city simulations can further be updated and changed as their physical equivalents change. These attributes of a DT, particularly its mirroring and ability to simulate factors, such as environmental conditions, population characteristics and movement, via mapping to dynamic real-world data, means that a place DT may not only allow more informed outbreak simulations by appropriate epidemiological models, but may also provide a significant solution to the cold start and privacy problems noted above that currently plagues the development of outbreak models [50]. Basing outbreak models on the demographic and locational foundations of city or place DTs can also further enhance the scope for repurposing such models effectively to incorporate the characteristics of diverse places and populations as well as provide the basis for adding new disease models for different pathogens [43].

Second, agent-based modelling and simulation of disease propagation has been shown to be a natural way to accommodate the effects of heterogenous characteristics and behaviors of individuals for simulating disease transmission and spread in a given geographic space [8, 40]. In particular, in an ABM-based disease simulation application, a disease transmission model is built on a generative and bottom-up process that can integrate three types of components: the agents and their susceptibilities and behaviors, the environment in which the agents operate by perceiving its state and acting accordingly, and finally, the mechanisms and processes that drive agent interactions [8, 32, 34]. Capturing the transitions involved in these interactions can further allow more realistic simulation of the outcomes of more specific and realistic interventions, such as imposing restrictions on specific types of businesses or other places, wearing of masks, and/or distribution of vaccinations, closely resembling those considered by public health officials.

These developments suggest that coupling disease ABM models with place DTs could provide a key tool for improving simulation of the propagation of pathogens in complex real-world settings [2, 5]. Here, we develop CitySEIRCast (Fig. 1), a multidimensional, multi-resolution, multi-scale DT simulation framework coupled with a flexible ABM and High-Performance Computing (HPC), as a modelling tool to more realistically simulate the transmission of a pandemic among the diverse subpopulations and settings of an urban locality, focusing on the COVID-19 pandemic as an example. Given the dynamic evolution of information regarding the epidemiology of SARS-CoV-2 and the shifting foci of interventions and priorities of city and county officials, we also focused on the adaptability of CitySEIRCast to produce outputs that are better able to reflect these changes over time. Although our co-simulation framework is applied in the context of the COVID-19 pandemic in Hillsborough County, Florida, a feature arising from our objective to make the system interoperable is designing CitySEIRCast in such a way as to make it easily adaptable to other cities and to other infectious diseases of concern.

**Fig. 1.**
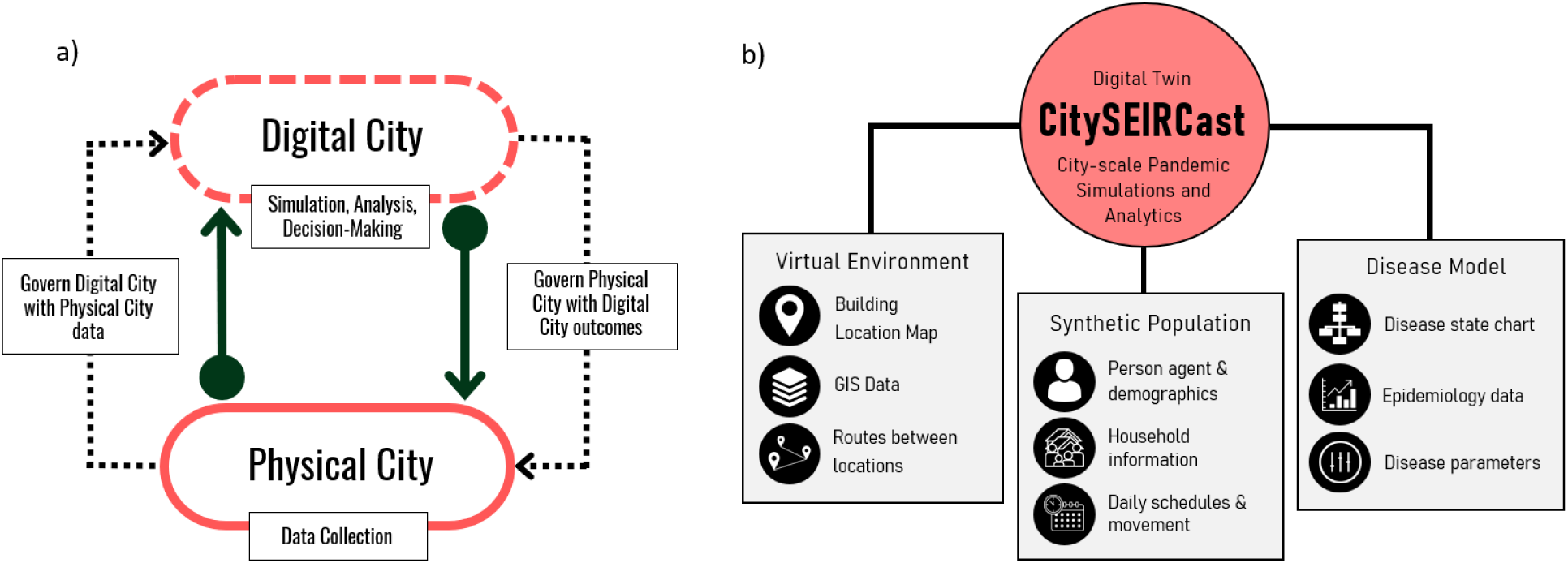
(a) A schematic representation of a digital city as a virtual twin of a physical city. (b) Our framework combining a digital city consisting of the virtual environment (buildings, routes) and synthetic population with the ABM disease model.

The paper proceeds as follows. In the next Section, we present the background details of ABMs and DTs with descriptions of their respective modelling frameworks. In Section 2, we describe the methodology, algorithms, workflows and data pipelines used to build CitySEIRCast. In Section 3, we present the main results of our coupled DT-ABM co-simulation system. In Section 4, we discuss the potential impact of our work for using coupled DT-ABMs as tools for modelling diseases in real-world societies and highlight the next stage of work required to leverage CitySEIRCast for co-simulating cities and urban infectious disease transmission dynamics.

### 1.1 Literature Review

In this section we discuss and compare existing tools and frameworks, related to DTs for health, including for COVID-19, and to agent-based disease modelling in general.

#### 1.1.1 The Digital Twin

The technological advancements of Industry 4.0—smart systems, artificial intelligence and machine learning, Internet of Things (IOT), big data management, cloud and edge computing, among others [57]—have piloted the development of the DT, a novel technology that merges the physical and digital worlds. The DT can be defined as a virtual model of a physical entity or process of any scale and all its interacting relevant components and properties [16, 25, 63]. The three main constituents of a DT, as established originally by Grieves [25], are the physical entity, the virtual replica of the physical entity, and the mutual communication between the physical entity and the virtual replica. Tao et al. [64] developed the five-dimension DT concept that includes data and services as two additional DT components. In this conceptual model six two-way connections exist between all the elements of the model - between physical entity and services, physical entity and data, physical entity and virtual world, virtual world and data, virtual world and services, and between services and data.

The virtual or digital replica must copy the physical world’s characteristics, elements, complexity, processes, interactions, external factors, and events with high fidelity and resolution, and to accomplish this, the DT includes and interacts with multiple interdisciplinary technologies, devices, and methods [16, 64]. There is thus no defined method or platform for creating the virtual twin [63]. Nevertheless, the method or platform chosen must have the ability to construct a virtual model of high accuracy and fine-grain detail, which can be verified by validation methods [63]. At the current state of DT technology, the physical entity to be replicated can be a device or machine or a city, a biological being (including human beings), a system, a system of systems, an environment, or a process. In the case of DT applications to public health, most employed thus far have focused on improving patient management and demand [56], although its potential for the prediction and management of infectious disease outbreaks in populations is increasingly receiving attention [2, 5, 50]. In the latter case, the virtual world needs human behavioral models that capture the expected actions of the inhabitants of the physical world and predict their actions under changing patterns. One way to do this is by adding an ABM layer [5], which generates ‘agents’ as entities with constraints to simulate expected real-life behaviors of the inhabitants residing in the physical world [32]. Rule-based models that drive the virtual agent’s ability to reason may also be needed [9].

Ultimately, the DT functions to aid in decision-making for improving the physical entity. The DT has the ability to test and analyze the outcomes of what-if scenarios and possible strategies to solve challenges before implementing them in the real world. With the seamless communication and transmission of data between the two worlds, the virtual space is constantly updated with data from the physical space and the physical space is informed with the results of what-if scenario testing in the virtual space. A feedback loop is formed (Figure 1a), where the physical world implements decisions based on the outcomes of different interventions in the virtual space, and over time the virtual space is updated with the outcomes of the interventions decided to be implemented in the physical space [53].

Constructing city DTs for supporting disease outbreak analytics and forecasting usually involves modeling the disease of interest and the heterogeneities involved, tracking cases or other health events attributed to the disease, and simulating individuals of the affected population as well as the complex social dynamics between themselves, with the surrounding environment, and with the infectious agent. However, there are a variety of methods by which DTs for disease have been created, which is a testament to the flexibility and adaptability of such frameworks.

Thus, Deren et al. [17] proposed the Smart City Public Epidemic Service System to integrate smart city simulation frameworks with healthcare institutions as well as patient health and movement data. Patient health data is used to assess the height and severity of the epidemic curve and movement data is used to determine the spatiotemporal trends of the epidemic. Using smart city infrastructure, the Epidemic Service System tracks, locates, and follows up on confirmed cases, and is therefore able to perform analyses of the dynamics of disease transmission, detect areas where risk of transmission is high, and send warnings of exposure risk to citizen smartphones. The system relays its epidemic analysis to government and health organizations to make in-tandem decisions regarding the strategies to adopt to halt disease transmission.

During the COVID-19 pandemic, DT frameworks specific to COVID-19 were proposed to help alleviate the impact of the pandemic. These DTs were constructed with different approaches and are tailored to fill gaps in the strategies to reduce the health and economic impact of the pandemic, but fundamentally they are all efforts to capture accurate COVID-19 disease transmission dynamics, pandemic trends, population behaviors during pandemic times, and the effectiveness of public health prevention and mitigation measures. Barat et al. [5] developed a DT framework to analyze the effect of non-pharmaceutical interventions to halt the spread of COVID-19 using the help of a DT of Pune City, India, coupled to an ABM. The ABM aspect of the Pune DT captures the demographic distribution of the population, their movements and interactions, and the strategic interventions that can be imposed by public health authorities. The authors program each type of agent with their own time schedule and movement pattern; and in the case of places, the type of agents that frequent the place and the type of agent interactions that can occur inside a place.

Pang et al. [50] outlined a multi-city COVID-19 project where each city and its population, spatial, and epidemiological aspects are modeled via DTs that are capable of self-improving by learning from itself and other city DTs through what the article calls local and global updates. Local updates refer to the collection and storage of historical and real-time public health data through IoT devices or relevant reliable sources of data. Global updates instead refer to cities uploading their parameters to a central server for other cities to learn and update their DTs. The federated updates are thought to be significant for helping authorities make public health decisions because a city that has taken a public health measure that others have not can upload its parameters before and after taking the measure so that other cities can conclude whether or not to impose the said measure.

Zhao et al. [73] proposed iGather, a smart contact tracing platform in the COVID-19 context that uses DT methods to precisely and anonymously map individuals and their real movement patterns through digital devices in a virtual space. The DT creates a link between individuals, community, workplaces, and health institutions to provide health guidance, alert individuals of possible exposure, track cases temporally and spatially, and identify areas of high transmission.

In contrast, and highlighting the diversity of DT construction methods, Meuser et al. [45] created a DT of a city in a game platform called ‘Cities: Skylines’ designed to build and manage highly detailed cities. The researchers modified the game platform to add an infectious disease dynamic layer to simulate COVID-19 transmission. A realistic population is replicated in this game by assigning demographic characteristics and movement patterns to the simulated individuals in order to capture accurate COVID-19 disease dynamics. Their framework may be adapted to other infectious diseases.

Pilati et al. [54] assembled a DT of COVID-19 mass vaccination centers to coordinate resources, reduce wait-times, and synchronize patient walk-ins while keeping measures to prevent COVID-19 transmission inside the facility. It achieves these goals by dividing the process of vaccination into phases, collecting time data of these phases at a case-study vaccination center with smartphones, and reproducing it in a virtual simulation of the vaccination center to digitally test different methods in which wait times can be reduced. With the DT tool, the researchers were able to find time-effective strategies specific to the situation of the vaccination center modeled.

With the cutting-edge developments of Industry 4.0 (Internet of Things (IoT), 5G, big data, blockchain, artificial intelligence, and machine learning) along with surging interest and investment in concepts such as cryptocurrencies and self-driving cars, a future of digital and smart cities may already be materializing [16]. The DT comes hand in hand with this trend of digitalization of city infrastructure. Given the interconnected nature of the DT and reliance on real-time capabilities, the smartness of a city facilitates the construction of a city DT and, vice versa, smart cities take benefit from city DTs for management of services. The data captured by IoT sensors and devices spread throughout a smart city to monitor the state and assess the needs of a city can be integrated in a DT for real-time updates, and in turn the DT can affect the smart cities through IoT broadcasting [16]. Some major cities are starting to invest resources to move towards a smart infrastructure and create their DT. Such is the case of Shanghai [70], New York City, starting with its Brooklyn Navy Yard [61], Singapore with project Virtual Singapore [59], and Helsinki with projects Helsinki 3D [11] and Kalasatama Digital Twins [36] for urban planning, sustainability, development, and other city-specific management.

#### 1.1.2 Agent-based Models

Agent-based models (ABMs) represent a computer simulation framework in which instead of using a single monolithic model, the dynamics of a system is captured through the actions of agents interacting with each other and with their environment [32, 41]. An agent is assigned different attributes that characterize it, such as behavior and types of interaction with environment components and other agents. The actions of agents are governed by a set of rules that direct their individual behaviors and interactions. As a result of this heterogeneity and stochasticity embedded in their individual behavior, ABMs can capture unexpected aggregate phenomena that result from the combined individual behaviors in a model [13]. This allows the use of ABMs for modelling complex social systems.

ABMs have gained popularity in the study of disease transmission due to their ability to capture complex human behavior and risk characteristics, contact networks, spatial hotspots, and other elements involved in disease transmission that standard mathematical models lack [32]. ABMs are able to capture realistic social interactions at a very fine scale by simulating attribute-dependent contact networks as well as social interactions inside different types of locations based on the schedule, structure, and dynamics of the location (i.e., workplaces, schools, long-term care facilities). An ABM designed to analyze disease transmission models individual agents that make up the population of interest by assigning attributes and behavior rule sets to each agent based on population data, and by developing a compartmental disease process model for the agents to navigate through. As the simulation progresses in this scheme, susceptible agents will come into defined contact with infectious agents and acquire a probability of getting infected with the disease. Infecteds can also become diseased or recover from infection and turn into immunes.

Similar to DTs, there is no defined methodology to follow for building an ABM which affords high levels of flexibility and adaptability in its construction. A variety of agent-based simulation frameworks have been developed recently to simulate and analyze disease transmission and public health interventions within the context of the COVID-19 pandemic [2, 5]. Here, we describe a selection of published work that demonstrate how ABMs can and have been used to investigate features of the complexity of COVID-19 transmission focused on interactions between heterogeneous populations, infection processes, contact networks, and control options.

Agrawal et al. [2] proposed an ABM to simulate and evaluate the effects of non-pharmaceutical interventions on the COVID-19 pandemic before imposing them on the public. The model features age-specific interactions, contact tracing and quarantine, as well as total and partial lockdowns, providing the means to model disease transmission dynamics under the highly heterogeneous conditions that typically govern contagion and response to epidemic spread in the real-world.

Similarly, Alagoz et al. [3] built COVAM, an ABM to analyze the impact of adherence to social distancing measures in New York City in the context of COVID-19. Imposing social distancing measures at different stages of the pandemic and at different levels were studied using a population modeled with realistic demographic attributes, contact networks, testing scenarios, and probability of cooperating with social distancing. The authors showed that the model was able to replicate the number of confirmed COVID-19 cases in the city.

Kerr et al. [34] developed Covasim, an ABM simulator to predict future patterns of the COVID-19 pandemic, test possible intervention strategies, and manage resources by modeling population demographics, social and transmission interactions that vary by types of locations (schools, hospitals, long-term care institutions, households), age-dependent patient outcomes, and disease parameters specific to a country or region. The simulator is completed with a variety of interventions, including social distancing, mask wearing, vaccination plans, testing of suspected symptomatic and asymptomatic cases, contact tracing, quarantine and isolation, among others. It is thought to achieve robust results by the capturing of these aspects that influence the spread of disease and control outcomes.

In the same vein, Chang et al. [10] presented an ABM to simulate the COVID-19 pandemic and its impact in Australia, calibrated to replicate real COVID-19 transmission dynamics. Several measures were tested with the model, including varying levels of isolation and quarantine of confirmed cases and contacts of cases, social distancing, closures of community spaces, and international travel regulations.

Ozik et al. [49] developed CityCOVID, an ABM simulator based on their existing social interaction computational platform, ChiSIM, applied to the Chicago area. The synthetic population statistically captures the demographic characteristics of the Chicago population. The synthetic agents are assigned hourly activities based on certain demographic characteristics, which guides movement from place to place to reproduce age- and place-specific social interactions and behaviors in the virtual space. The synthetic agents become exposed to COVID-19 while interacting with infectious agents in the simulation.

On the other hand, Singh et al. [60] adapted EpiGraph, an existing epidemic simulator, and used it to evaluate control measures and lockdown scenarios to halt the spread of COVID-19 in Madrid, Spain. An ABM layer was added to capture realistic social behavior and networking dynamics of the individuals that make up the real Madrid population as well as to model movement within and out of the city.

Hinch et al. [30] built a COVID-19 ABM simulator called OpenABM-Covid19 as a tool for policymakers and other stakeholders in order to use simulations of the COVID-19 pandemic with regards to making management decisions on optimal interventions and preventative measures. The simulator captures region-specific demographics and age-dependent social interactions to assess non-pharmaceutical interventions, contact tracing, and vaccination plans.

Finally, Suryawanshi et al. [62] proposed an ABM of the city of Kolkata, India, focusing on spatial and contact components of COVID-19 transmission. The synthetic agents are characterized by age, income, presence of comorbidity, workplace, family, and places they frequently visit. This model also analyzes different possible public health interventions for executing in the city across different levels and probabilities of compliance.

### 1.2 COUPLING CITY DIGITAL TWIN WITH ABMs: CitySEIRCast

The sections above indicate that coupling a City DT with ABMs that more realistically mimic the processes of infection transmission can allow the development of a co-simulation system to better address the complexities of disease transmission in an urban population [5, 16]. Thus, while the city DT can allow a faithful rendering of the properties that characterize an urban location and its population, the ABM can allow the simulation of disease transmission dynamics that takes a fuller account of the spatio-temporal multidimensional risk factors that may drive pathogen transmission. Here, we describe the development of CitySEIRCast, a multidimensional, multi-resolution and multi-scale DT-ABM co-simulation framework, to evaluate the ability of such a modelling construct to analyze and predict the dynamics of disease outbreaks, including emergence, propagation and infection persistence, using the transmission of COVID-19 in Hillsborough County, Florida, as a case study.

## 2 FRAMEWORK AND METHODOLOGY

Our DT is divided into three main modules (Fig. 1(b)). The first module is the virtual environment of the setting under study, constructed using city information data that maps and categorizes buildings, routes between places, and environmental conditions [58]. The second module is the synthetic population, which is a statistical representation of the corresponding population demographics, household information, the daily schedules, and routes of each synthetic person. The third module is the disease model layer, which combines the states and the parameters and processes governing the transmission of a disease as well as the epidemiological data specific to the modeled region. The subsections that follow detail the computational workflows, components, and data pipelines of each module and their application in the context of the spatio-temporal progression of the COVID-19 pandemic in Hillsborough County, Florida.

### 2.1 Virtual Environment Module

The virtual environment refers to the digital reconstruction of Hillsborough County, which involves precisely mapping and classifying every building and location type as well as street layouts present in the County. We applied the City Information Modelling (CIM) paradigm [14] to collect and weave these data from a variety of sources to create a 3-D model of the urban environment representative of the County. While CIM can combine both below and above ground structures, here we focused on above ground structures only. We extracted and used building information contained with the parcels data from the Hillsborough County Property Appraiser (https://downloads.hcpafl.org/) [28] as well as data on zip code boundaries to perform this construction. The parcels data come in the form of shapefiles and provide information on several features of the buildings in the county, including location, use type, size, built levels, and number of rooms. The 3-D virtual city model is then constructed by integrating the building information above within a geographic information system (specifically QGIS) using the algorithm displayed in Fig. 2. Specifically, this is performed firstly by reading the county zip code boundaries provided in a GeoJSON file accessed from [27] into QGIS. Then, we extract and add building location (latitude and longitude) and building type data, categorized into households, schools, workplaces, and community places (which include shopping malls and retail stores, grocery stores, places of worship, and outdoors) from the parcels data [28]. Polygonal geometry of buildings and street layouts/routes are then added from OpenStreetMap (https://www.openstreetmap.org/) [12] to complete the city modelling. Fig. 3 shows a comparison of the 3-D virtual representation of a portion of Hillsborough County created using our algorithm against the satellite view [65] for that portion, which indicates that the physical environment of the area in question is replicated well by the CIM.

**Fig. 2.**
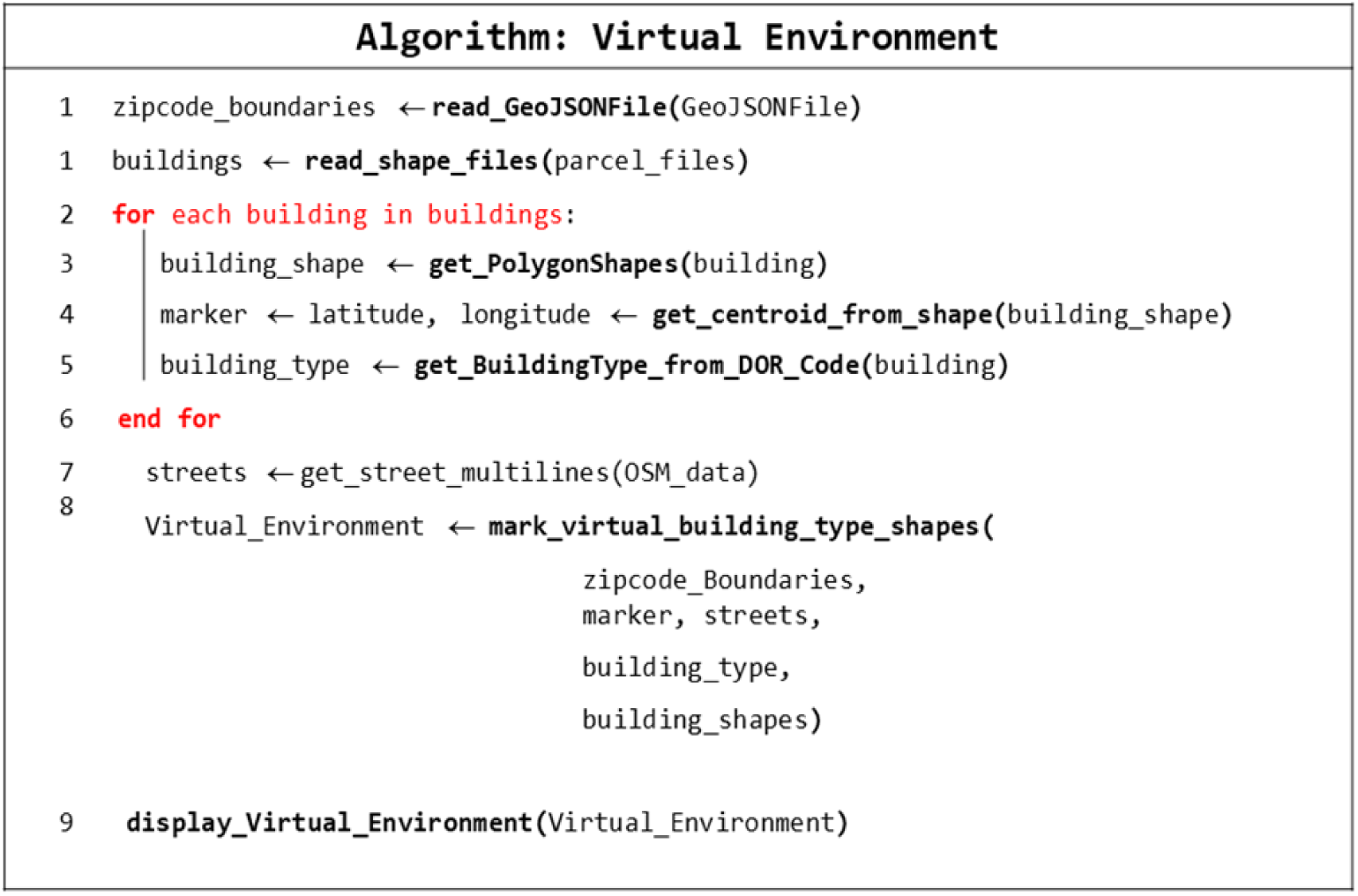
Algorithm to generate the building types that make up the virtual environment of the DT.

**Fig. 3.**
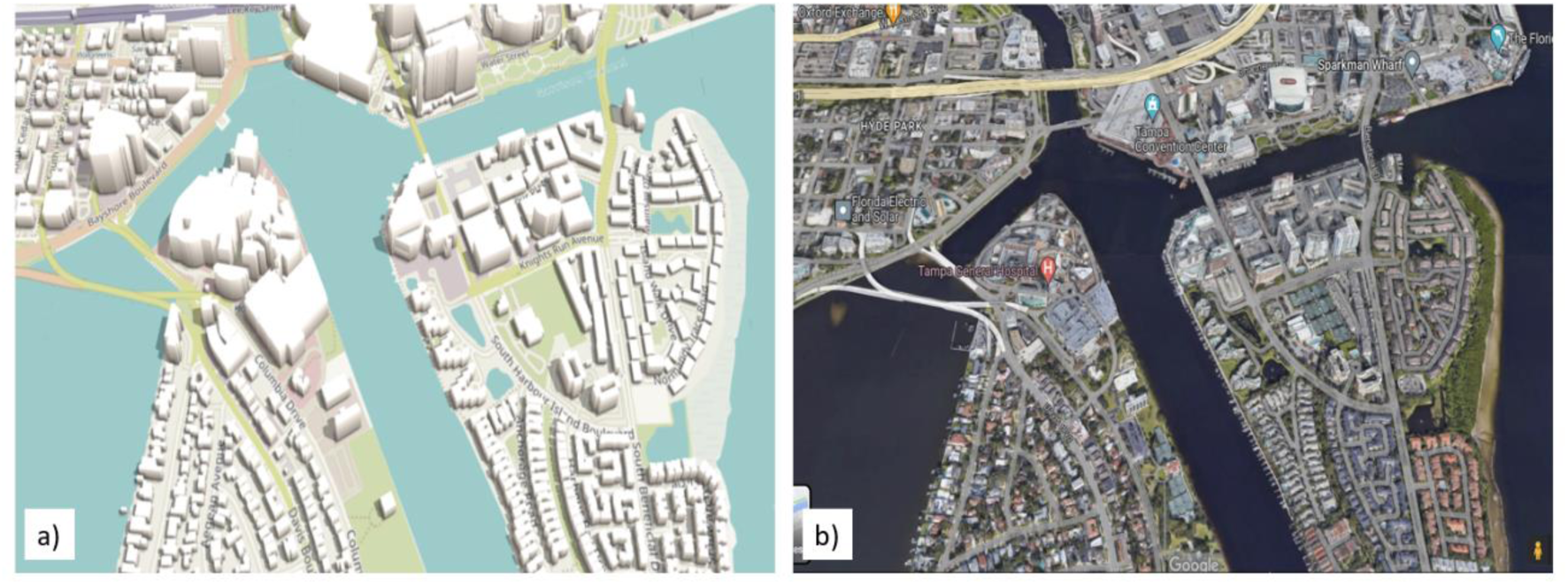
(a) A portion of Hillsborough County showing 3D building locations, shapes, and sizes. (b) A satellite image of the same area in Hillsborough County.

### 2.2 Synthetic Population Module

The synthetic population module generates a population of agents that is representative of the demographics of the region of interest, in this case Hillsborough County and its 52 zip codes, based on census data available from PolicyMap© (www.policymap.org) [1]. The specific objective is to generate the individual synthetic agents and then locate them to households according to household size. These agents are assigned demographic attributes, including age, gender, race, ethnicity, income, and mobility patterns, based on the population characteristics found for zip code (see example for zip code number 33612 shown in Table 1) and the origin-destination (OD) matrix describing movement patterns for individuals using the algorithm shown in Fig. 4. We collected data on demographic attributes at the zip code level, and the household and school sizes within the county, to serve as inputs for executing this algorithm. The module generates the synthetic population through the following steps (Fig 4).

**Fig. 4.**
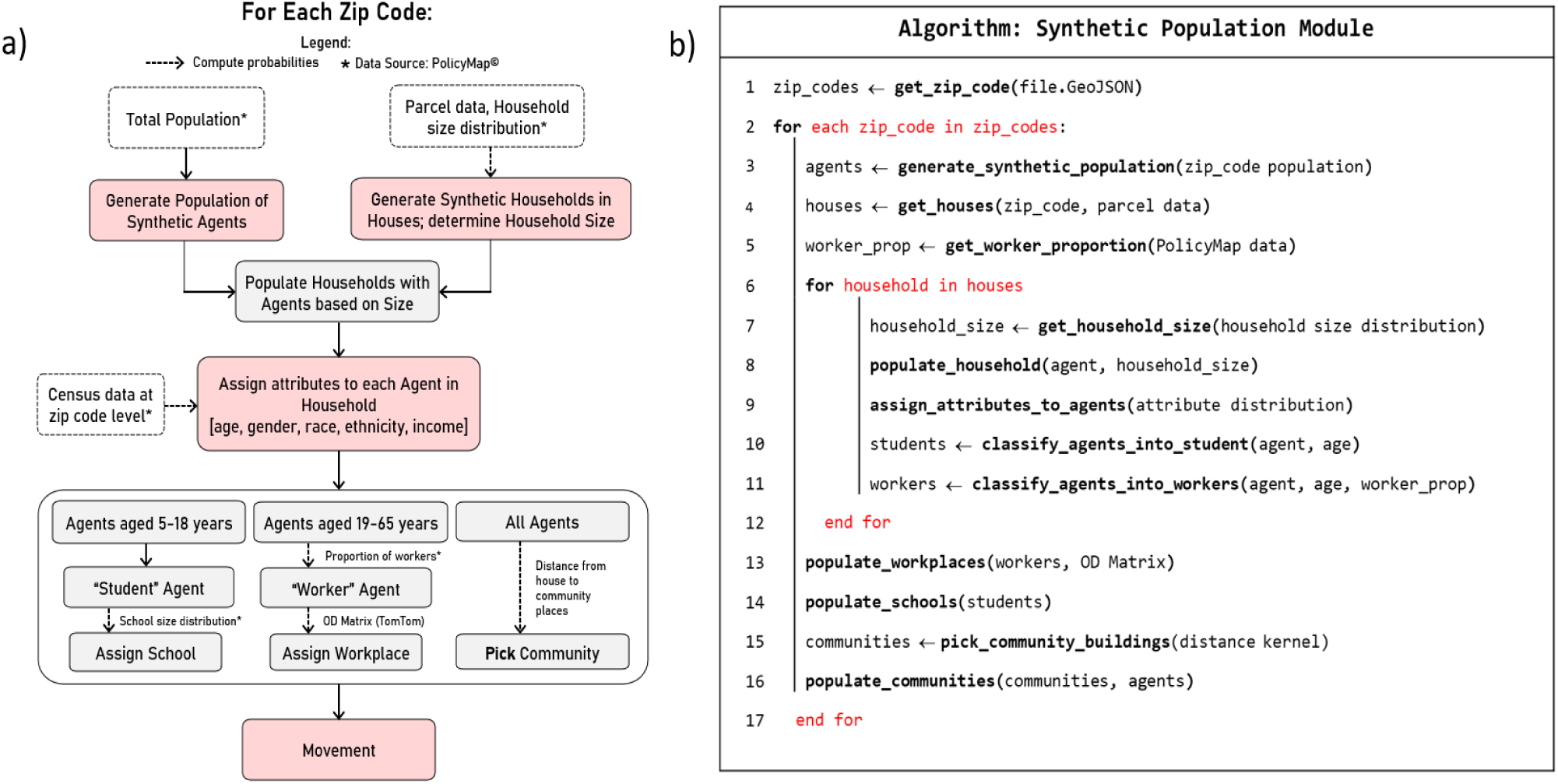
The synthetic population generation module. a) Flowchart of the steps taken by the module to generate the synthetic population. b) Details of the synthetic population generation algorithm.

**Table 1.** Summary statistics for the population residing in zip code 33612 based on census data obtained from PolicyMap [1]. Similar data for all zip codes were used to construct the synthetic population of Hillsborough County.

| Population Statistics for Zip Code 33612 in Hillsborough County |  |
| --- | --- |
| Population | Count |
| Population size | 51745 |
| Age | Percentage (%) |
| Persons under 18 years | 21 |
| Persons between 18 and 64 years | 64 |
| Persons above 65 years | 12 |
| Race | Percentage (%) |
| White | 36 |
| African American | 30 |
| Asian | 3 |
| Native Hawaiian and other Pacific Islander | 0.1 |
| American Indian or Alaskan Native | 0.8 |
| Other Race | 15 |
| Two or more race | 15 |
| Ethnicity | Percentage (%) |
| Hispanic | 38 |
| Non-Hispanic | 62 |
| Gender | Percentage (%) |
| Male | 50 |
| Female | 50 |
| Income (\$) | Percentage (%) |
| 1 - 24,999 | 53 |
| 25,000 - 49,999 | 29 |
| 50,000 - 74,999 | 9 |
| 75,000 - 99,999 | 4 |
| 100,000 - 199,999 | 4 |
| 200,000 > | 1 |

#### 1) Generate a population of agents at the zip code level

The first step in the module is to generate a population of agents representing the population size for each zip code. We used the total number of people residing in a zip code based on the population data available from PolicyMap to accomplish this task [1].

#### 2) Generate synthetic households

Next, we create synthetic households for buildings labeled as a house or any type of residence in the parcel data [28]. The locations of these households are mapped based on their spatial coordinates as provided in the parcel data. Household size distribution in a zip code is constructed based on the number of households in a zip code, with size (single occupancy, 2-6 and 7+ occupants) allocated randomly based on the size distribution observed in each zip code.

#### 3) Assigning attributes to agents

We perform this by first randomly distributing agents to houses (which include single-family homes, apartments, and other types of residential buildings) based on household size (Fig 4). Demographic attributes are then assigned probabilistically to each agent viz. age, gender, race, ethnicity, and income, based on their actual distributions in each zip code. A sample of the resulting spatial distribution of races is shown in Fig. 5. A validation of synthetic population distributions against the available demographics data is shown in Fig. 6. This figure indicates that the synthetic population generated via the proposed algorithm closely mimics the real population in the county.

**Fig. 5.**
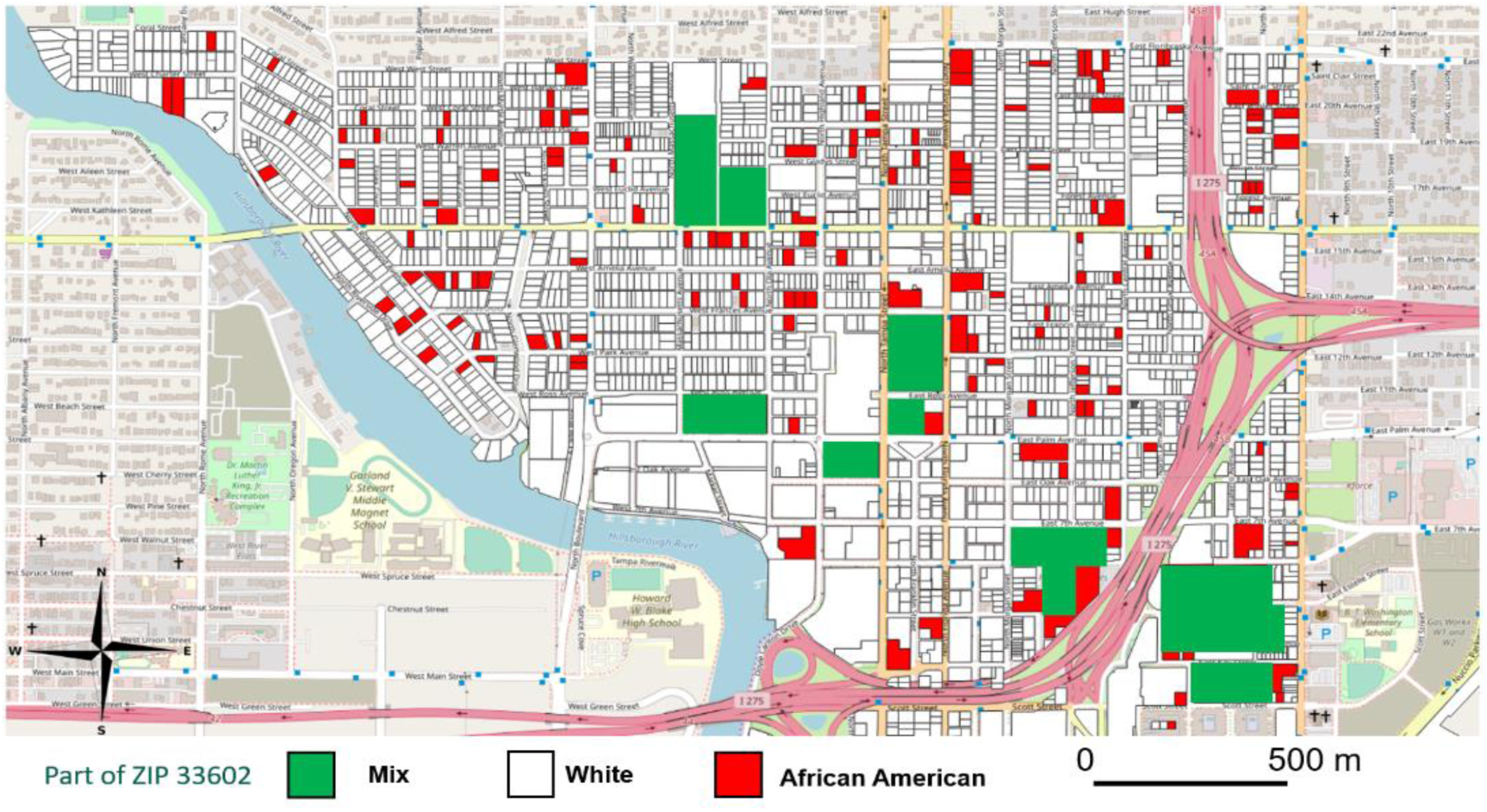
Distribution of races by in the digital twin. A part of zip code 33602 is shown. Buildings with African American, White and Mixed-Race population are shown in red, white and green respectively.

**Fig. 6.**
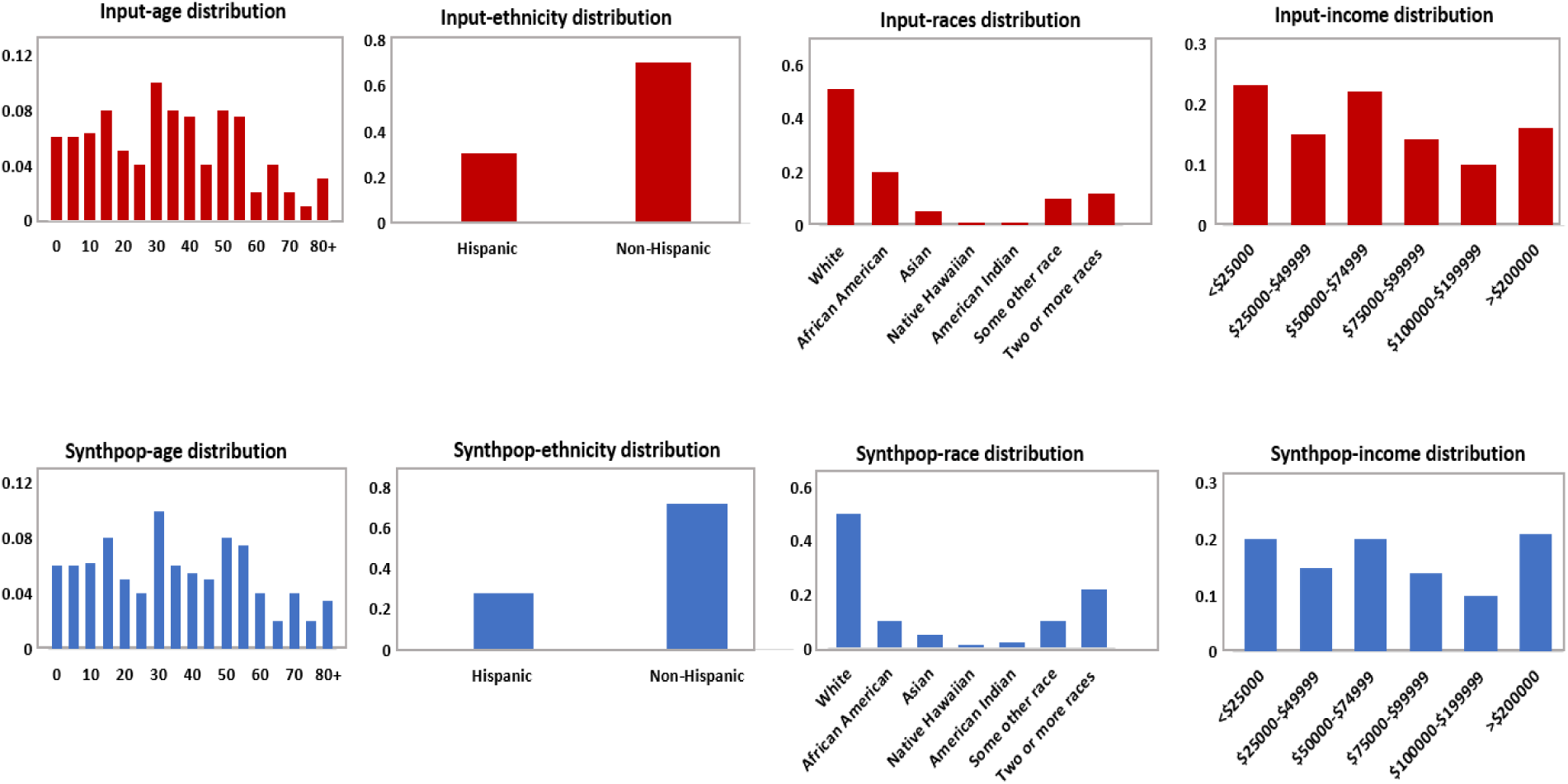
Validation of the synthetic population distributions (Blue) vs. actual distributions (Red) by age, race, ethnicity, and income.

Since the movement pattern of the agents are largely determined by their age and employment, we introduce the following categorization in the synthetic population model. Agents aged 5 to 18 are students who go to school, whereas agents from ages 18 to 65 are workers who go to workplaces, with the rest assumed to be unemployed. The probability of an agent belonging to each of these categories is in accordance with the proportions of each group of individuals residing in the zip code [1]. Each school-going agent is randomly assigned a school in the zip code of residence based on the capacity of the school, which is defined using the school size distribution of the county [1], and the number of school-going agents residing in the zip code. For the working agents, the zip code of their workplace is determined based on the OD Matrix constructed using mobility data (see below) provided by TomTom [68]. Essentially, this matrix gives the proportion of workers moving to their place of employment from one zip code to another or within the same zip code. Once the zip code of the workplace is decided for an agent, a random workplace is chosen in that zip code and assigned to that agent. A similar assignment of schools is performed for children who have to move outside their home zip codes for schooling. These categories and respective school/workplace assignment influence the mobility pattern of the agents, which we address next.

#### 4) Agent Movement

We used an OD matrix to inform the mobility patterns of agents in our city DT. An OD matrix is a data structure that represents the movement of people, goods, or vehicles between different locations. It is a table that shows the number of trips between each origin and destination pair, usually, at the minimum over a given time period. The rows represent the origins (O), while the columns represent the destinations (D). The cells in the table provide the number of trips between each origin-destination pair [39]. OD datasets can contain details of trips between two geographic points or, more commonly, zones (which are often represented by a zone centroid). The latter combined with the total number of trips from an origin to destination zones allows calculations of the fractions of trips made by people within a county versus the fraction of these trips made to other counties.

We used TomTom’s OD analysis engine [68] to estimate the OD matrix for informing the mobility of agents in the DT. TomTom OD analysis is based on real time Floating Car Data (FCD) obtained by combining signals aggregated monthly from anonymous GPS enabled cars and mobile phones [67]. We provided zip code boundaries to the TomTom OD analysis platform in order to output an OD matrix representing travels or people movements within and between zip codes. This information is used to assign workplaces and schools to the fractions of employed adults and school-going individuals both within their zip codes of origin and to other destination zip codes (see above).

We also additionally calculate the cumulative distances travelled by individuals based on the OD distributions by estimating the distance kernel applicable to each zip code. This presents the typical distances people travel based on their location of origin, and is used to model movement to community spaces, as the movement of people to community areas is more complicated compared to workplaces and schools. For example, people may visit different community areas at different points of the day, and there are a wide variety of community places to be visited. To accommodate this heterogeneity, we use the determined distance kernels to assign weights for travelling to communities by an agent residing in a zip code.

Finally, as the movement algorithm used by the ABM simulates the motion of agents in the virtual environment through a simulation time step of 6 hours, we further divided each day into four time-segments (morning, afternoon, evening and night) and estimated the OD matrix/distance kernel for each time period. This allows us to simulate the daily activity of agents according to the time of the day as well as when different categories of buildings are open at different times of the day. For instance, schools are open from mornings to afternoons. Similarly, less people go to communities and workplaces after midnight (Fig. 7). In the ABM, we model this feature by sending students to schools during selected segments of the day, with similar restrictions placed on other age categories with regard to work (as well as for pursuing leisure activities).

**Fig. 7.**
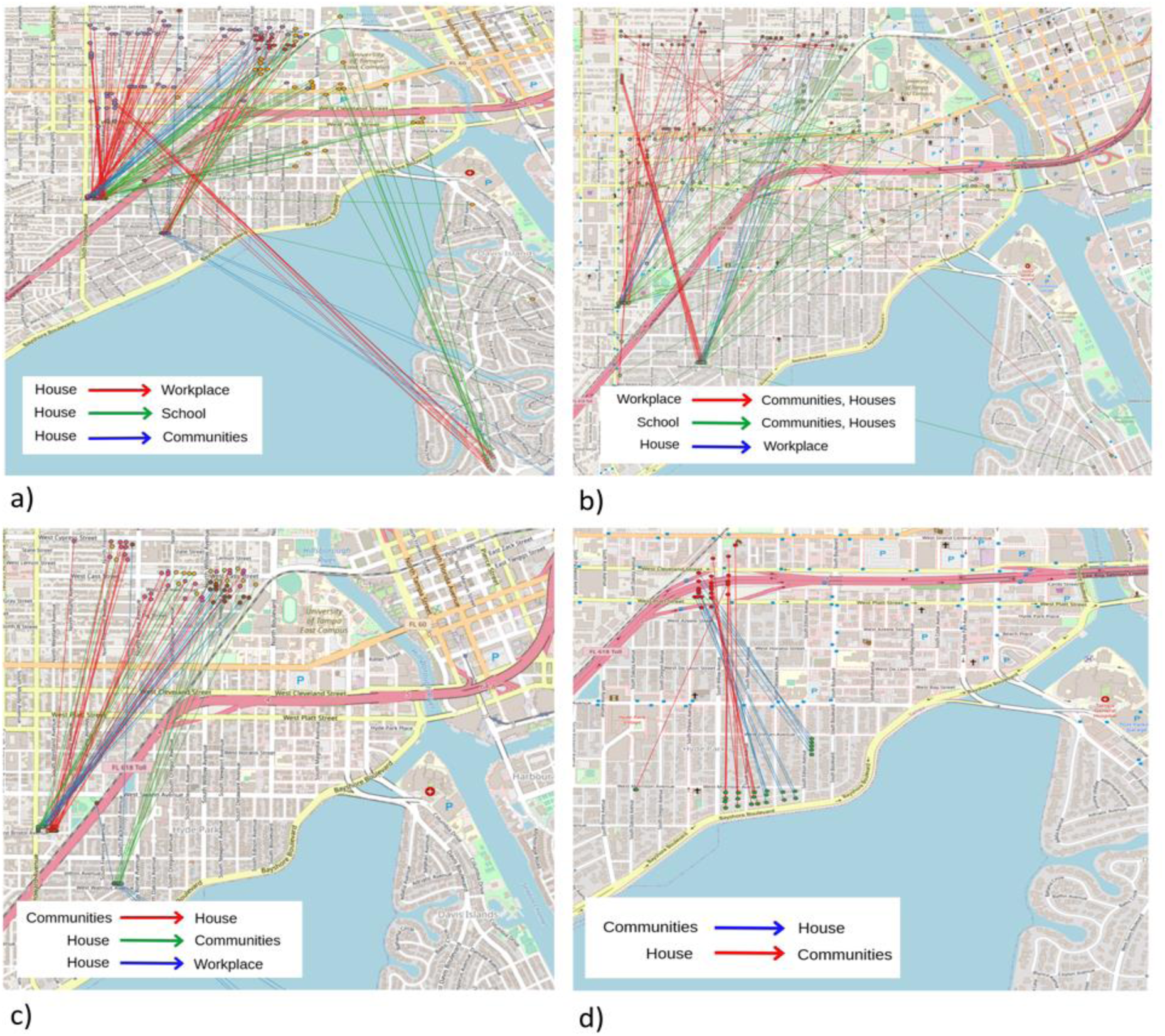
Agent movement from and to different places during the 6-hour time blocks. a) 7am to 12pm, b) 1pm to 6pm, c) 7pm to 12am, d) 12am to 6am.

### 2.3 Disease Module

#### 2.3.1 The ABM model

The ABM disease model, illustrated in Figs. 8 and 9, follows the synthetic agents as they move through different disease stages. Susceptible individuals are exposed and become infected by coming in contact with infectious individuals. Exposed and infected individuals can then fall into pre-symptomatic or asymptomatic categories. Asymptomatics recover after a mean period of 4 days. Individuals in the pre-symptomatic category initially do not show any symptoms for about 2 to 6 days and then proceed to show mild symptoms. After approximately 7 days, the individual either recovers or shows severe symptoms and may require hospitalization. Once hospitalized, individuals can recover or deteriorate further and require critical care. Individuals in critical care either recover or die. The risk of developing severe symptoms, requiring critical care, or death is age dependent (See Table S1 in the supplementary document). Age dependence in later stages of the infection also serves as a proxy for comorbidities, which are also age-dependent in general. All recovered individuals can become susceptible again after a period ranging from 6 months to 1 year. We model the effects of vaccines, along with mask-wearing and social distancing, as control measures. The vaccination strategy at the time of writing for COVID-19 initially consisted of two doses, the second given within 3 to 4 weeks’ time after the first, followed by a waning period of five months, and then 1st booster is given at about the 6 months mark. Thereafter the individual can have the second booster after waiting for another six months. Individuals with different levels of immunity induced via vaccines can also become exposed and follow a similar infection route as fully susceptible, albeit with a lower chance of developing severe disease and hence requiring critical care. The effect of mask-wearing is modeled as reducing the chance of exposure to the virus by variable degrees as does social distancing. In our model, we implemented a time-varying mask-wearing and social distancing behavior based on COVID-19 search patterns in Google trend data [24], as described in Section 2.3.3. In the initial part of the pandemic, lockdowns with varying levels of restrictions on workplaces, schools, and communities over a period of 3 to 4 months were also modeled to mimic the scenario followed in Hillsborough County.

**Fig. 8.**
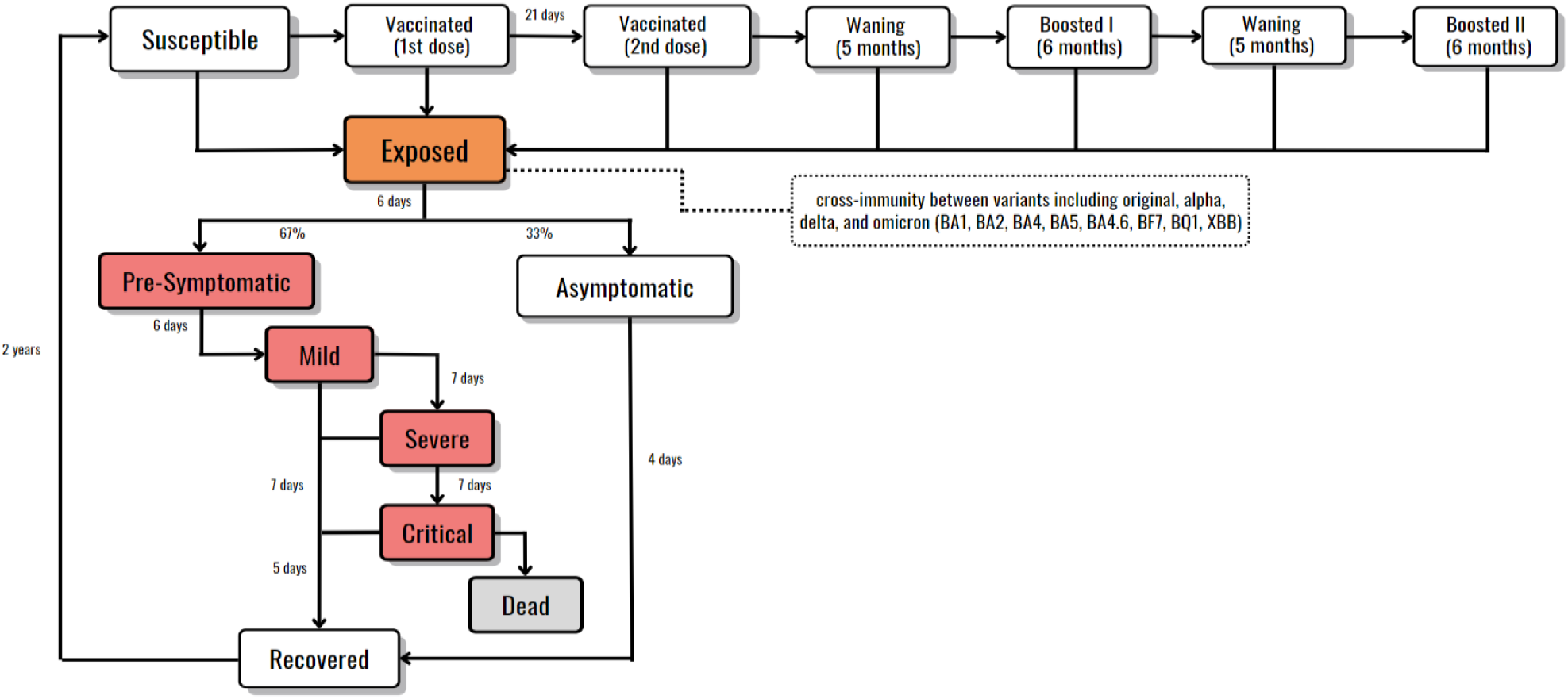
Disease progression model including variants and vaccine doses.

**Fig. 9.**
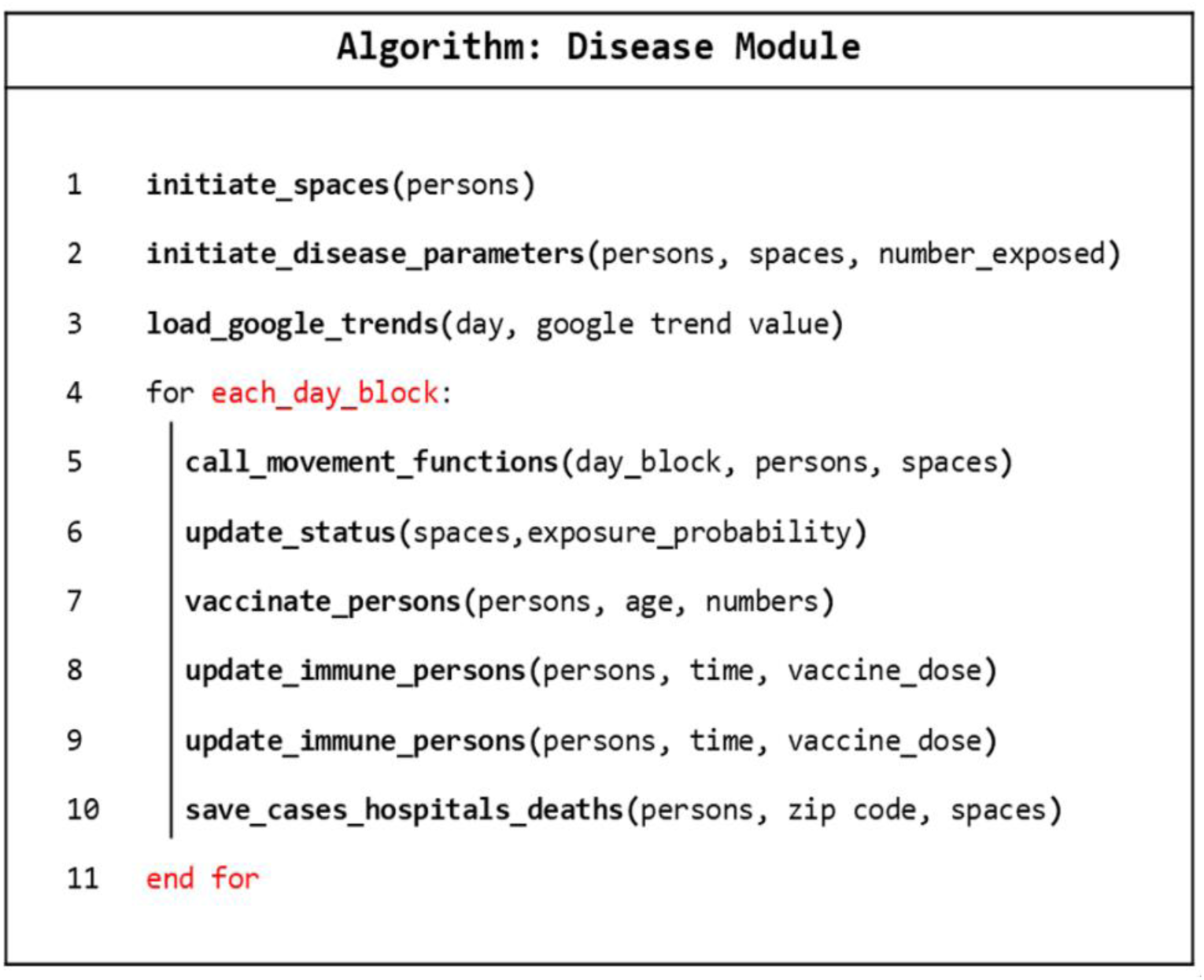
Disease transmission algorithm followed by the CitySEIRCast simulator.

We model the spread of infection in the simulator following the works of Ferguson et al. [21] and Agrawal et al. [2], extending the model to include all the variants in Hillsborough County over time and human behavior changes as estimated via Google trends time series data [24]. At each time window, a force of infection is computed for each individual *n* based on proximity to other individuals in different spaces (home, neighborhood, workplace/school, local community, or a random community) following the time schedules followed by individuals: as an example, schools are active only during the first two quarters of the day. Furthermore, an individual experiences force of infection from the prevailing strains in these spaces; the individual picks up each strain randomly with a probability of picking a strain depending on the force of infection it produces for the individual. Human protective behavior *H^b^*(*t*) in different spaces is modeled based on Google trends search data, *gt*(*t*), reflecting social curiosity about COVID-19 over time [24]. An individual is compliant and takes measures including social distancing and mask wearing based on *gt*(*t*) and prevailing conditions at different spaces, viz whether workplaces are open or not. At home these measures would reflect washing hands regularly and practicing isolations/quarantine in the case of infected family members. At time *t*, each susceptible individual transits to the exposed state with probability 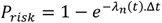 (Fig. 10), where Δ*t* is the simulation time step taken to be 6 hours, and *λ_n_*(*t*) is the force of infection (which a function of all the rules above) experienced by the n*^th^* individual interacting with other individuals across different spaces (Fig. 10).

**Fig. 10.**
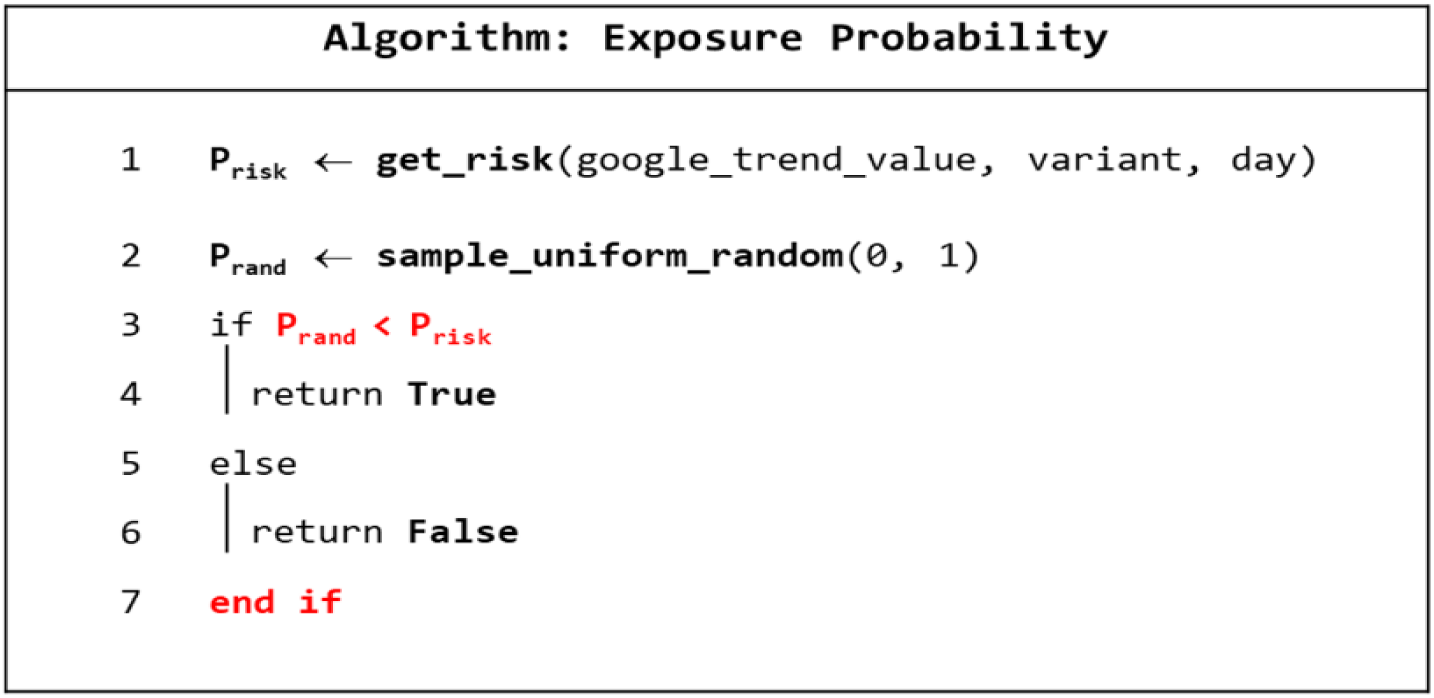
Exposure probability algorithm followed by our CitySEIRCast simulator.

The explicit form of the force of infection is given as

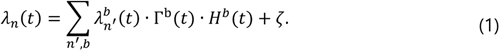

Here *b* is the set of building types that an individual visits and *n^′^* is the set of individuals the agent interacts with during the visit. The summation term represents the force of infection when the agent in is specific buildings (houses *h*, neighbors’ houses *h^nbr^*, schools *s*, classrooms *s^c^*, workplace *w*, or project groups *w^p^*). The exact expression for 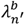 depends on the building type:

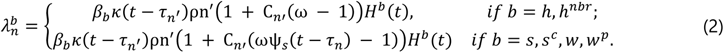

When the agent is in transit *T*, community area *c*, or random community area *rc*, the interaction between the agents follows a different functional form given by ζ,

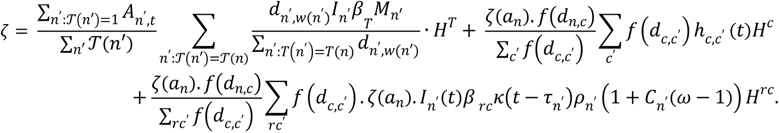

where,

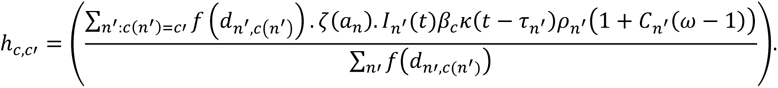

Here *I_n_*_(*t*)_ = {0, 1} is based on whether an agent is infectious or not, *ρ_n_* reflects an individual’s variability in relative infectiousness (coming from a gamma distribution) and *C_n_* = {0, 1} indicates whether an agent is severely infected and hence more infectious by a factor of (ω − 1). By contrast, the force of infection is reduced by a factor, (Ψ*_s_*_,*w*_(t − τ*_n_*)ω − 1), in the case of work and school absenteeism due to severe infection resulting from exposure at time τ*_n_*. The severity factor and absenteeism are taken to be ω = 2 and Ψ*_s_*_,*w*_(t − τ*_n_*) = {0.1, 0.5} [2]. The individual sees a relative travel related contact rate ζ(*a_n′_*) specific to age, we take this to be 0.1, 0.25, 0.5, 0.75, 1, 1, 1, 1, 1,1, 1, 1, 0.75, 0.5, 0.25, and 0.1 for the various age groups in steps of 5 years, with the last one being the 80+ category. Infection-stage-related infectiousness is taken to be κ(t − τ*_n_*) at time *t*. For the disease progression described in the previous section, κ(t − τ*_n_*) is 1 in the pre-symptomatic and asymptomatic stages, 1.5 in the symptomatic, hospitalized, and critical stages, and 0 in the other stages. Individuals using public transport transmit more if they travel longer distances, while visits to communities in other zip codes contribute to the force of infection proportional to the distance kernel function *f*(*d_c_*_,*c*_) (see Tables 2 and 3).

**Table 2.** Parameter table.

| Parameter | Interpretation | Prior distribution/Value |
| --- | --- | --- |
| $\beta_h$ | Transmission rate in home settings | U(0.1, 0.2) |
| $\beta_h^{nbr}$ | Transmission rate in home settings | U(0.1, 0.25) |
| $\beta_w$ | Transmission rate in work settings | U(0.2, 0.3) |
| $\beta_{wp}$ | Transmission rate at work in project | U(0.1, 0.3) |
| $\beta_c$ | Transmission rate in community settings | U(0.1, 0.25) |
| $\beta_{rc}$ | Transmission rate in random community | U(0.1, 0.25) |
| $\beta_s$ | Transmission rate in school settings | U(0.3, 0.5) |
| $\beta_{sc}$ | Transmission rate at school in classroom setting | U(0.1, 0.2) |
| $\beta_T$ | Transmission rate in public transport. | U(0,0.05) |
| Gama distribution shape parameter $\theta$ | Shape parameter for gamma distribution for immunity | 2 |
| Mean natural Immunity period $\gamma_0$ | Rate of joining susceptible class after recovering | 2 years (mean) |
| Mean Immunity period vaccinated 1<sup>st</sup> dose $\gamma_1$ | Rate of joining susceptible class after recovering due to vaccine 1 | 1 month (mean) |
| Mean Immunity period vaccinated 1<sup>st</sup> dose $\gamma_2$ | Rate of joining susceptible class after recovering | (mean) |
| Mean Immunity period vaccinated 1<sup>st</sup> dose $\gamma_3$ | Rate of joining susceptible class after recovering | 2 years (mean) |
| Mean Immunity period vaccinated 1<sup>st</sup> dose $\gamma_4$ | Rate of joining susceptible class after recovering | 2 years (mean) |
| Immunity period (natural or vaccine induced) | Individual agents rate of joining susceptible class after recovering | Gamma($\frac{\gamma_i}{\theta}, \theta$) |
| Mean incubation period $\sigma$ | Exposed to pre-symptomatic/symptomatic period | 4.5 days |
| $\sigma_i$ | Individual incubation period | Gamma($\frac{\sigma_i}{\theta}, \theta$) |
| Pre-symptomatic fraction | Fraction of individuals that would eventually be symptomatic | 0.67 |

The force of infection described above along with the transition probabilities in Table 2 and Table S1 of the supplementary document are used to transition the agents through the disease stages as described.

#### 2.3.2 Arrival of new strains

We started the simulator with the original strain of COVID-19 virus. That is, at the start of the simulation 100 percent of infectious individuals are infected by the original strain. Subsequently, as new variants are reported in the real world (see Table 3 for the time of arrival of variants), we introduce these strains in the simulation as follows. On the day of the first reports, a small fraction of infectious agents is infected by the new variant. In the simulation, this new variant is represented by a new set of transmission, hospitalization, and death rates. As time progresses the proportion of agents infected with the new strain increases until the new variant overtakes the existing strain. This is to be expected as the new variant is, in general, more transmissible than its existing counterpart. This competition between the strains is reflected by the gradual increase in the effective force of infection experienced by the agents from the new strain.

**Table 3.** Functional forms and miscellaneous variables.

| Function/variable | Interpretation | Functional form/value |
| --- | --- | --- |
| $f(d)$ | Distance kernel | $\frac{1}{\left(1 + \left(\frac{d}{a}\right)^b\right)}$<br>a=20.751, b=3.384 |
| $T_{\Omega 0}$ | Household transmission rate scales as $n^{1-\alpha}$ | 0.8 |
| $r_{\alpha}$ | Relative transmissibility of alpha variant | 1.3 |
| $r_d$ | Relative transmissibility of Delta variant | 3 |
| $r_{\Omega}$ | Relative transmissibility of Omicron variant | 1.2 |
| $T_{\alpha}$ | Time of arrival of alpha variant in Hillsborough | 30<sup>th</sup> December 2020 |
| $T_d$ | Time of arrival of Delta variant in Hillsborough | 25<sup>th</sup> April 2021 |
| $T_{\Omega 0}$ | Time of arrival of Omicron variant in Hillsborough | 2<sup>nd</sup> Dec 2021 |
| $T_{\Omega 1}$ | Time of arrival of Omicron sub-variant in Hillsborough | 23<sup>rd</sup> Jan 2022 |
| $T_{\Omega 2}$ | Time of arrival of Omicron sub-variant in Hillsborough | 8<sup>th</sup> May 2022 |
| $T_{\Omega 3}$ | Time of arrival of Omicron sub-variant in Hillsborough | 8<sup>th</sup> May 2022 |
| $T_{\Omega 4}$ | Time of arrival of Omicron sub-variant in Hillsborough | 17<sup>th</sup> July 2022 |
| $T_{\Omega 5}$ | Time of arrival of Omicron sub-variant in Hillsborough | 28<sup>th</sup> Aug 2022 |
| $T_{\Omega 6}$ | Time of arrival of Omicron sub-variant in Hillsborough | 2<sup>nd</sup> Sep 2022 |
| $T_{\Omega 7}$ | Time of arrival of Omicron sub-variant in Hillsborough | 12<sup>th</sup> Sep 2022 |
| Vaccine effectiveness 1<sup>st</sup> dose | Effectiveness of vaccine in inducing immunity to individuals after 1<sup>st</sup> vaccine dose | 0.75 |
| Vaccine effectiveness 2<sup>nd</sup> dose | Effectiveness of vaccine in inducing immunity to individuals after 2<sup>nd</sup> vaccine dose | 0.95 |
| Vaccine effectiveness 3<sup>rd</sup> dose | Effectiveness of vaccine in inducing immunity to individuals after third vaccine dose | 0.99 |
| Vaccine effectiveness 4<sup>th</sup> dose | Effectiveness of vaccine in inducing immunity to individuals after 4<sup>th</sup> vaccine dose | 0.99 |

#### 2.3.3 Google Trends and human behavior

We use Google trends data [24] for COVID-19-related search terms to model human protective behavior. This model assumes that social media interest in a particular topic can translate to a behavioral response by an agent. In the present case, we consider that interest in COVID-19 related search terms by agents would lead to higher compliance with intervention measures, and hence in a reduction in the force of infection. The degree of reduction in the force of infection 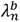 in space *b* based on individuals’ compliance status is obtained via a Bernoulli trial *B*(1, *p*), where the probability of compliance is *p* = *gt*(*t*). Noncompliant individuals see a relatively higher force of infection in a given space whereas compliant individuals experience a reduced force of infection in that space (Equation 1). Additionally, during different phases of the pandemic, the high non-compliance observed for community places (shops, malls) is implemented by ignoring Google trend values in this space, while maintaining the limited restrictions followed in workplaces and schools. This was specifically the case during the delta and the subsequent omicron transmission surges [19]. To allow for uncertainty in human behavior, we consider ensembles of Google trends data, created by adding and subtracting 10 percent to the original data and time-shifting by two weeks. The full set of Google trends and members of the ensemble, as described, are shown in Fig. 11.

**Fig. 11.**
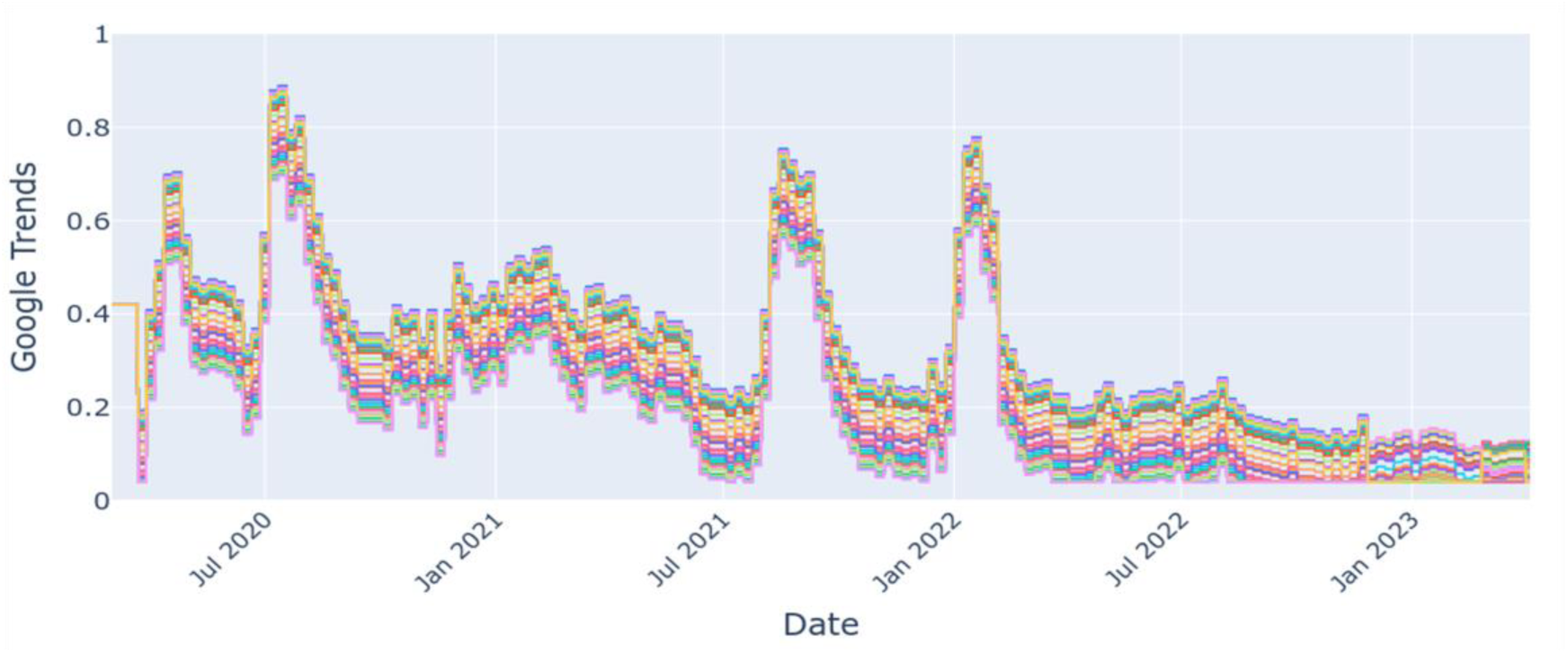
Google trends ensemble generated through addition of 10% variance to the original trend in the queries made. In total, we derived 100 ensembles, 50 of which sampled from the actual trends data and another 50 by shifting the full time series to the right by two weeks. The data are used as a proxy for mask compliance as well as determine the degree of exposure in places/spaces where people interact, viz. home, school, community, and workplace.

#### 2.3.4 Lockdown mandates in Hillsborough County, Florida

The lockdown and closure mandates in the State of Florida or by Hillsborough County authorities are implemented in the simulator following the schedules available from Hillsborough County mandate documents. To accurately model the timing, span, and extent of lockdowns, we referred to Executive Orders (EO) enacted by governing bodies where they detail each measure [18, 19]. Guidelines regarding the closure of schools were obtained from the Florida Department of Education and Hillsborough County Public Schools [22, 23]. Lockdowns were imposed in several phases, i.e., Phase 1, Phase 2, Phase 3, and finally the release of all lockdowns as part of the “Safe. Smart. Step-by-Step” recovery plan devised by [55]. Phase 0 refers to the initial measures taken from March 9, 2020, when the State of Florida declared COVID-19 as a public health emergency, which included encouraging all state residents to stay at home except for essential services, while restaurants and public schools were closed. Phase 1, implemented on April 29, 2020, continued to encourage staying at home, and allowed restaurants and retail stores to open at 25% of indoor capacity. Phase 1 was extended on May 15, 2020, with the modification that restaurants and retail stores could operate at 50% of indoor capacity. Phase 2, implemented on June 5, 2020, allowed all non-essential businesses, including retail stores, to fully reopen, while restaurants remained operating at 50% indoor capacity. Phase 3, implemented on September 25, 2020, continued on Phase 2 regulations but allowed restaurants to operate at no more than 50% indoor capacity. Essential businesses, grocery stores, financial institutions, and places of worship remained open throughout all phases while public schools and universities were operated entirely remotely. On May 3, 2021, the State of Florida ordered that all COVID-19-related orders and mandates be lifted across the state [19]. The lockdown and phase timeline are depicted in Fig. S1 and Table S3.

The level of closures of various spaces/buildings is incorporated in the simulator via the parameter Γ^b^(*t*) where *b* corresponds to spaces categorized into schools, workplaces, community spaces.

#### 2.3.5 Vaccination schedules

We implement the same vaccination schedule as that followed in Hillsborough County. The first doses of vaccine were delivered in January 2021. Following the first dose, individuals were required to have a second dose to complete the vaccination sequence. Subsequently, the first booster doses were available. During the simulations, the Hillsborough-wide daily vaccination data by dose was approximated by polynomial curves shown in Fig. S2. Vaccine efficacy is implemented according to dosage viz. first dose is 75% efficient, while second dose has a 95% efficacy and booster has a 99% efficacy, and a waning individual has a reduced efficacy of 90% between second dose and subsequent booster [48]. The total number of vaccines on a given day were distributed by age and that distribution changed over time according to vaccination guidelines [18–20, 46], see Table S4 in the supplementary document.

### 2.4 Parameter Calibration

We use a Bayesian calibration technique to learn values of the subset of parameters related to rates of transmission across different spaces (Table 2), based on initial sensitivity analysis. Starting from a uniform prior distribution assigned to these free parameters, we obtain a posterior distribution of parameters by minimizing the root mean square error between simulated and reported daily case data [29]. As the time series extended, we refined the parameters as the pandemic unfolded and new data regarding variants and new control measures, including mask-wearing, lockdown, and vaccinations became available. Specifically, when a new variant arrives the transmission, hospitalization and death factors are scaled according to variant properties (Table 2). Schematically the process of obtaining posterior distributions from data is shown in Fig. 12. Additionally, we assign each agent a natural and vaccine-induced immunity period obtained from a gamma distribution. The details of the priors used for estimating the transmission rates and the shape and scale parameters of the gamma distributions associated with the immunity states, and the other fixed parameters are given in Table 2. The time shift for Google trends ensembles is also treated as a parameter and is varied from 0 weeks to 4 weeks for the fitting procedure. The root-mean square error criterion between observed and simulated daily cases data for identifying the best fitting parameters at the county level is given by:

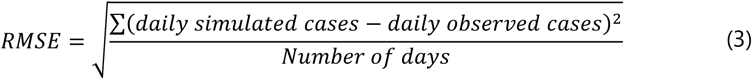

**Fig. 12.**
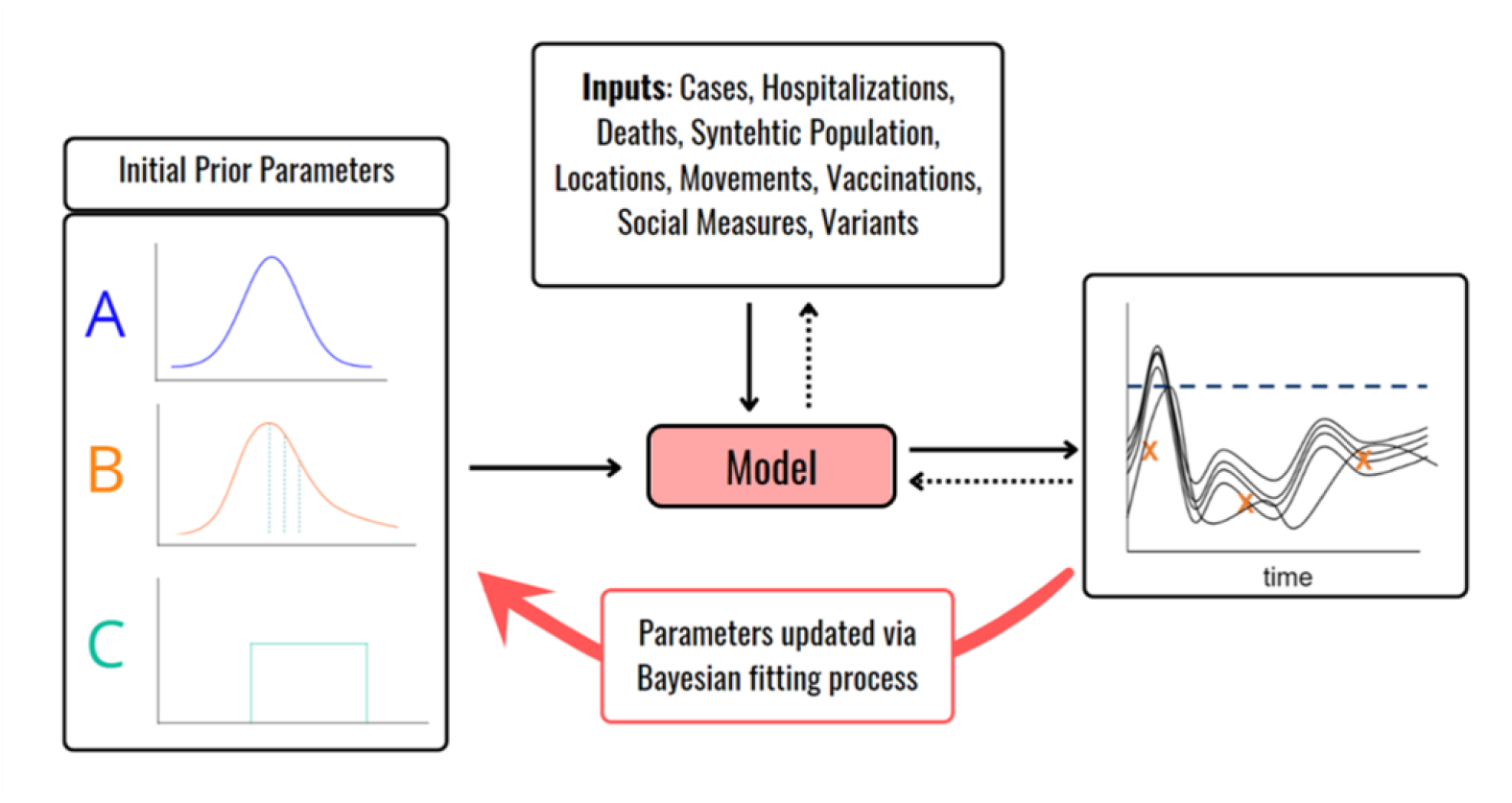
The Bayesian melding framework for parameter calibration using observed data. We used daily reported cases at the county level to calibrate the parameters listed in Tables 2 and 3.

We used this procedure to identify 200 best-fitting models that exhibited the lowest RMSE values, and deployed this ensemble to forecast the spatio-temporal propagation of COVID-19 in Hillsborough County.

### 2.5 Computation

As the DT is a bottom-up simulation, it starts with several types of input files: epidemiological data that includes case, admission, death, and vaccination data; population data that includes distributions of race, age, gender, income, and employment; and spatial data that includes locational and mobility data. These input files come together in the ABM layer, which uses the synthetic population, movements and disease status to simulate disease propagation. As noted above, the synthetic population, composed of agents that are combined with demographics and disease status, also needs to be distributed to the zip codes of Hillsborough County in a way that is statistically representative of the population of each zip code. Further, we also require simulating the detailed movement of individuals between zip codes to capture the reality of county residents going to work, school, stores, and other types of buildings outside of their zip code. These operations mean that both the DT simulation as well as the model simulations need to be computed in parallel and managed effectively for post-processing. Our solution is to harness the power of multiple CPU-cores and the larger memory capabilities of HPC to efficiently carry out these simulations on hundreds of cores.

#### 2.5.1 High-Performance Computing Architecture

To enable large-scale simulations in a distributed environment, we created an MPI cluster on Microsoft Azure, consisting of multiple virtual machines (VMs). The cluster leveraged 16 Azure VM HB instances, each with 96 cores and 384 GB RAM, resulting in a total of 1536 cores and 6.144 TB RAM across all VMs. This configuration ensured efficient parallel processing for the simulations, allowing the making of 1 month-ahead forecasts using a calibrated ensemble of 200 models to be completed over 3 days. Additionally, a Network File System (NFS) file share of 10 TB was used for data storage and sharing across the VMs. Initially, this was cloud-based which was then moved off cloud for improving speed of access to the data for visualization purposes. The HPC architecture is summarized in Fig. 13.

**Fig. 13.**
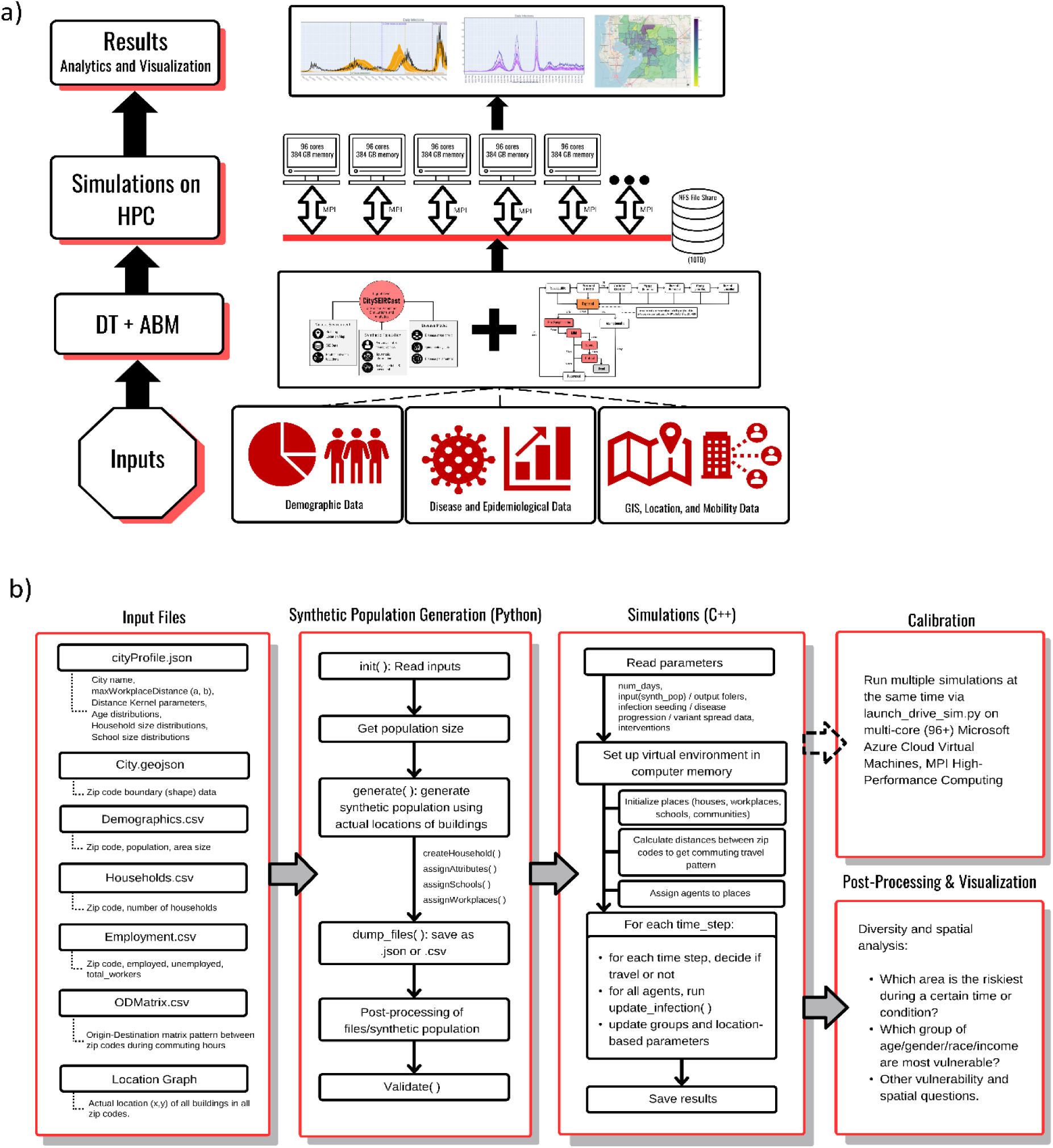
High performance computational framework for achieving the full calibration of CitySEIRCast simulator.

#### 2.5.2 Hybrid Python and C++ Execution

The workflow was further designed to incorporate both Python and C++ components for efficient parallel execution. Python was used to implement the MPI functionality for performing the ABM ensemble calibration, while the simulator itself was written in C++. This hybrid approach allowed seamless integration of Python’s high-level MPI capabilities with the performance advantage of C++ for executing the simulator. Each MPI process runs an instance of the C++ simulator. We split our 15,000 ensemble runs into batches of 96×16 (corresponding to number of cores in a single VM and the number VM’s as described in the previous section). This allowed for the calibration and forecasting computations to be carried out in a time-efficient manner.

#### 2.5.3 Workflow and resource management

The heterogeneous nature of our workflow, which incorporated Python, C++, and MPI components, required careful coordination and resource management. The high-level Python MPI functionality was used to distribute the simulation parameters across the VMs and to coordinate the execution of C++ simulator instances. This approach ensured efficient utilization of the available hardware resources and allowed for rapid switching between the different components. The overall workflow consisted of generating parameters in Python, distributing them to the C++/MPI simulations, and registering progress through database operations. This coordination was performed using MPI and Python’s in-built functionality, without relying on external workflow systems. The chosen approach allowed for effective management of in-memory user libraries and third-party packages, making it suitable for our complex simulation requirements.

## 3 SIMULATION RESULTS

In this section, we present the simulation results for the COVID-19 case study, focusing specifically on the spatio-temporal propagation of cases, hospitalizations and deaths at the county and zip code levels in Hillsborough County. We also show results for the effects of heterogeneous transmission among sub-populations, synchronization of cases across zip codes, and spatial patterns of immunity evolution across the County and its impact on the future transmission of the pandemic.

### 3.1 County-level analysis

We initiated the simulation of the pandemic with a seeding of exposed cases based on the zip code level cases reported in March 2020. We started with 10,000 sets of parameter vectors for the ABM model, with the values for each parameter drawn randomly from the prior distributions described in Table 2 using the Latin Hypercube sampling process. 200 best-fitting models are then selected for further simulation using the Bayesian calibration approach described in Methods. Fig. 14 shows the results of the county-level simulations for daily cases, hospitalizations, and deaths) from March 2020 to end of July 2023. The solid red, green and grey lines in the figure depict the mean values of the daily-cases, hospitalizations, and deaths respectively, whereas the shaded regions show the respective the 95% confidence intervals for each mean. The solid black line in each figure panel represents the actual numbers reported for daily cases, hospitalizations and deaths by the Department of Health, Hillsborough County. While the model predictions are able to capture the temporal wave-like patterns observed in the reported data for each clinical state reasonably well, it is apparent that the quantitative performance of the model contrasted with the data observed at different phases of the pandemic, particularly in the case of hospitalizations and deaths. We consider that this mismatch between predictions and observed data is a function of several factors, including parameter uncertainty that is not fully constrained by calibration to only daily case data, the consideration of only the Hillsborough population ignoring impacts of visitors, and problems with reporting of data, particularly in the case of deaths. Other issues could be related with uncertainties with regards to the actual implementation and compliance with the various social measures mandated by the authorities, the arrival times of the variants, and the movement model only approximately capturing the mobility of agents throughout the pandemic. This indicates that further investigation of parameter sensitivity, agent movements using better real-time data, agent compliance behavior, and inclusion of a data model for addressing errors in reporting, will be required to correct these anomalies. We are currently addressing the resolution of these issues to improve predictive performance.

**Fig. 14.**
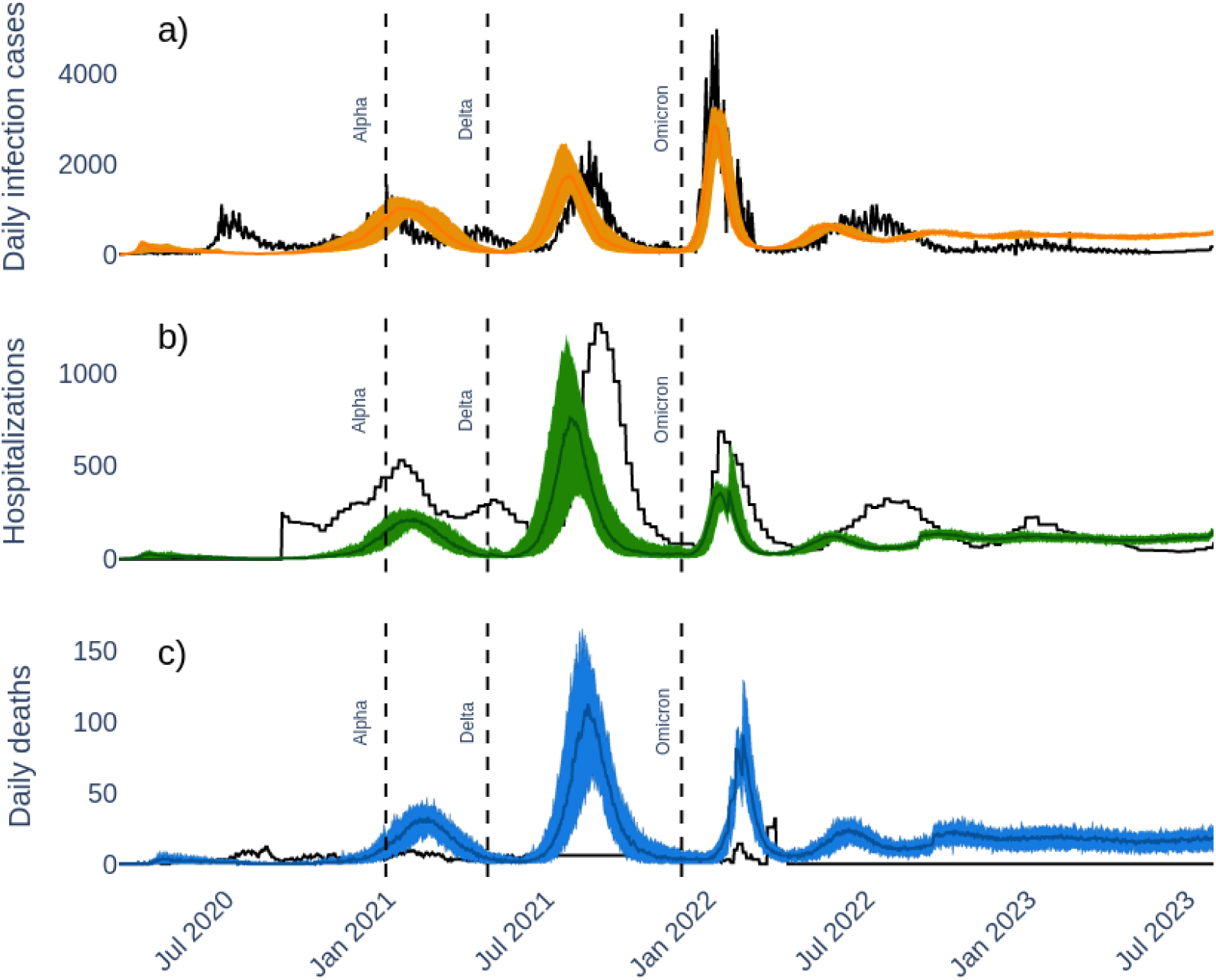
Daily infected cases, hospitalizations and deaths across the Hillsborough County. The shaded blue region indicates the 99 percent confidence interval obtained from ensemble of simulations using parameter priors as in Table 1. An ensemble of 10,000 sets of parameters from the prior distribution were simulated and a selection criterion by RMSE resulted in 159 best fitting (shaded region) curves against reported daily infection cases (black line). Vertical dashed lines reflect the dates of arrival of the major virus variants.

#### 3.1.1 County-level analysis by population sub-groups

A feature of the DT-ABM is that it allows sub-group analysis to be carried out for identifying and characterizing the most-risky population categories for a disease. Fig.15 illustrates the absolute daily case numbers predicted for the population of Hillsborough, divided by age groups, races, ethnicity, and income categories. These results show, on the one hand, that working age groups are the main contributors to the predicted daily cases across the county, whereas White people have the highest number of cases among all races. Similarly, lower-income groups contributed the most to the overall daily cases. On the other hand, an analysis based on proportions relative to population size did not reveal much difference in daily cases, even though hospitalizations and deaths were disproportionately high in African American and other minority groups, including Hispanics (data not shown).

**Fig. 15.**
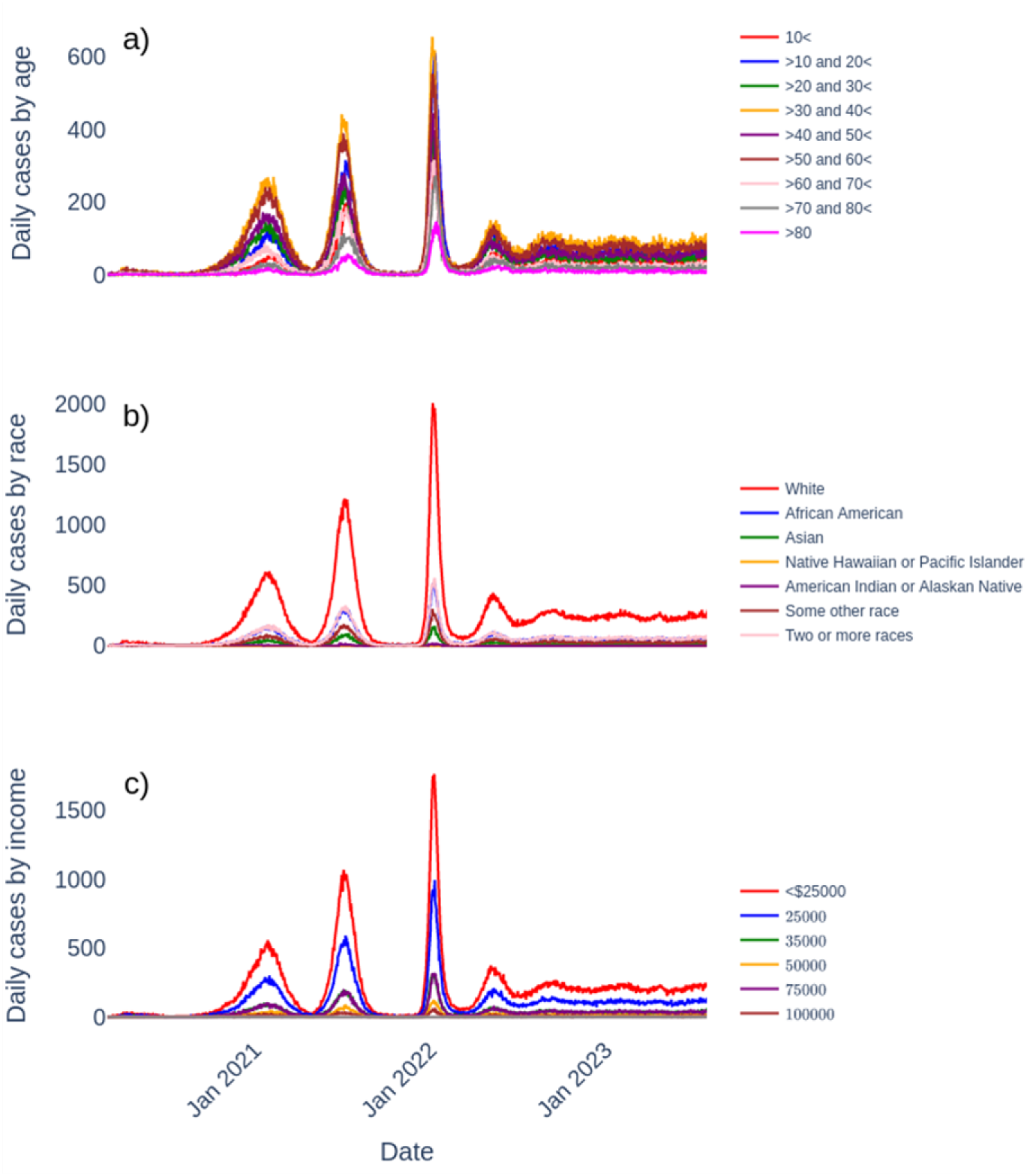
Daily infected cases by diversity groups (a) age, (b) race and (c) income.

### 3.2 Zip code-level analysis

This was performed on a subset of 5 zip codes chosen to represent different locations, population sizes, and infection patterns in the county. The analysis based on these zip codes reveals two key patterns. Firstly, some of these representative zip codes (and by extension some zip codes in the entire county) always show higher numbers of daily cases relative to the median cases predicted across all zip codes, while the numbers predicted in others are always below the median, as can be seen in Fig. 16. The grey shaded region in the figure indicates the 95% confidence interval constructed at each time for the combined predictions for all the 52 zip codes modelled, while the red curves show the corresponding median daily cases arising from these predictions. The dashed black lines show the simulated daily cases in the selected zip code and indicate each zip code’s infection levels relative to the median cases for all zip codes. These results demonstrate that throughout the pandemic, some of the 5 zip codes were consistently at higher risk for infection compared to the rest. This pattern can also be observed in the maps showing the spatial distribution of the cases predicted at the zip code level (Fig. 17). As can be seen, significant heterogeneity or spatial asynchrony between zip codes emerged early during the pandemic, which persisted over time (high infection zip codes remaining relatively highly infected), although there is also an interesting indication of changes in spatial synchrony across the zip codes.

**Fig. 16.**
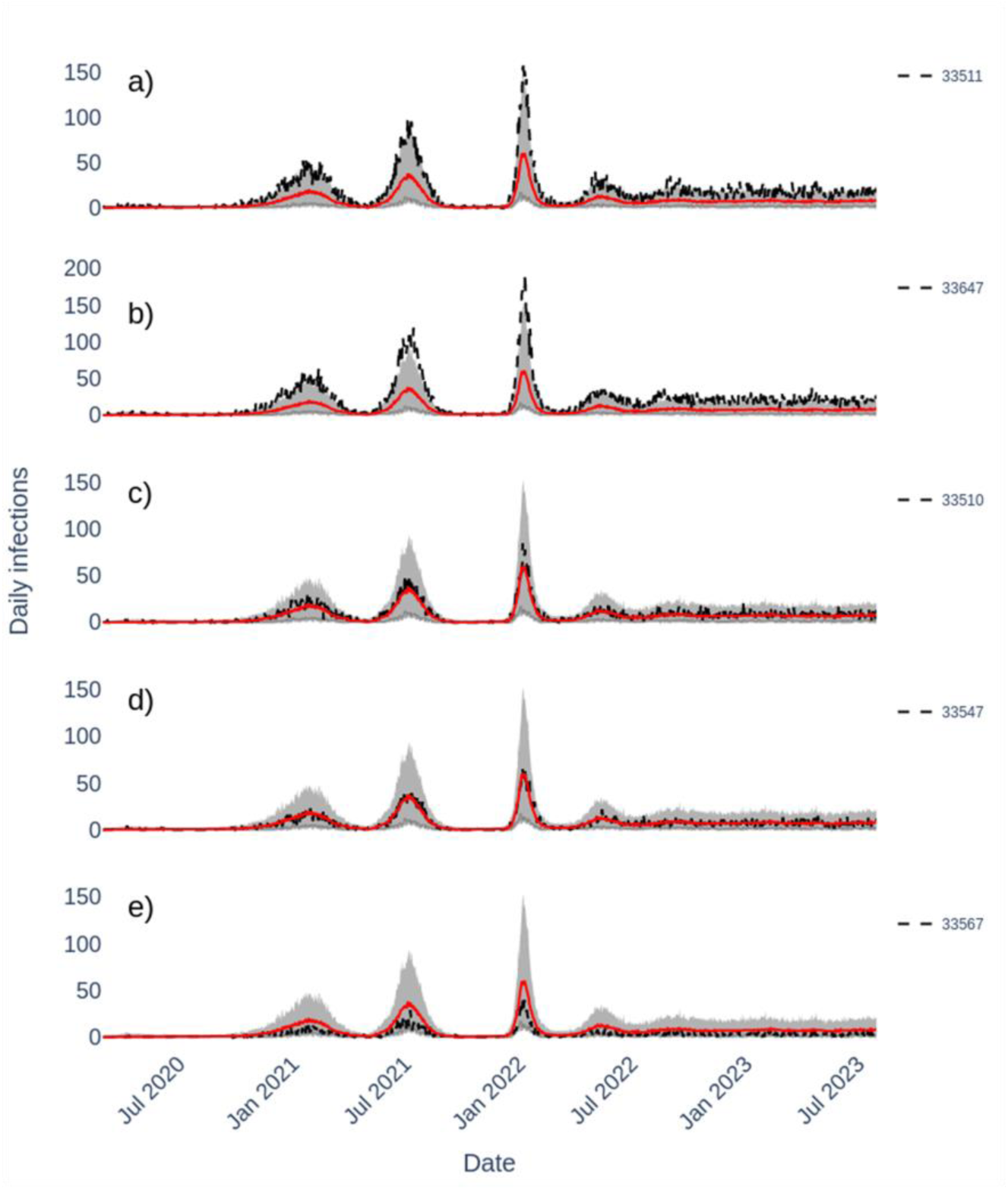
Daily infected cases in selected zip codes (see legends) indicating the zip codes relative position to the median cases (red line) over time across all zip codes in Hillsborough County. The grey shaded region indicates the 95% confidence interval constructed at each time for the combined predictions for all the 52 zip codes The dashed black curves correspond to the simulated daily cases for the selected zip code.

**Fig. 17.**
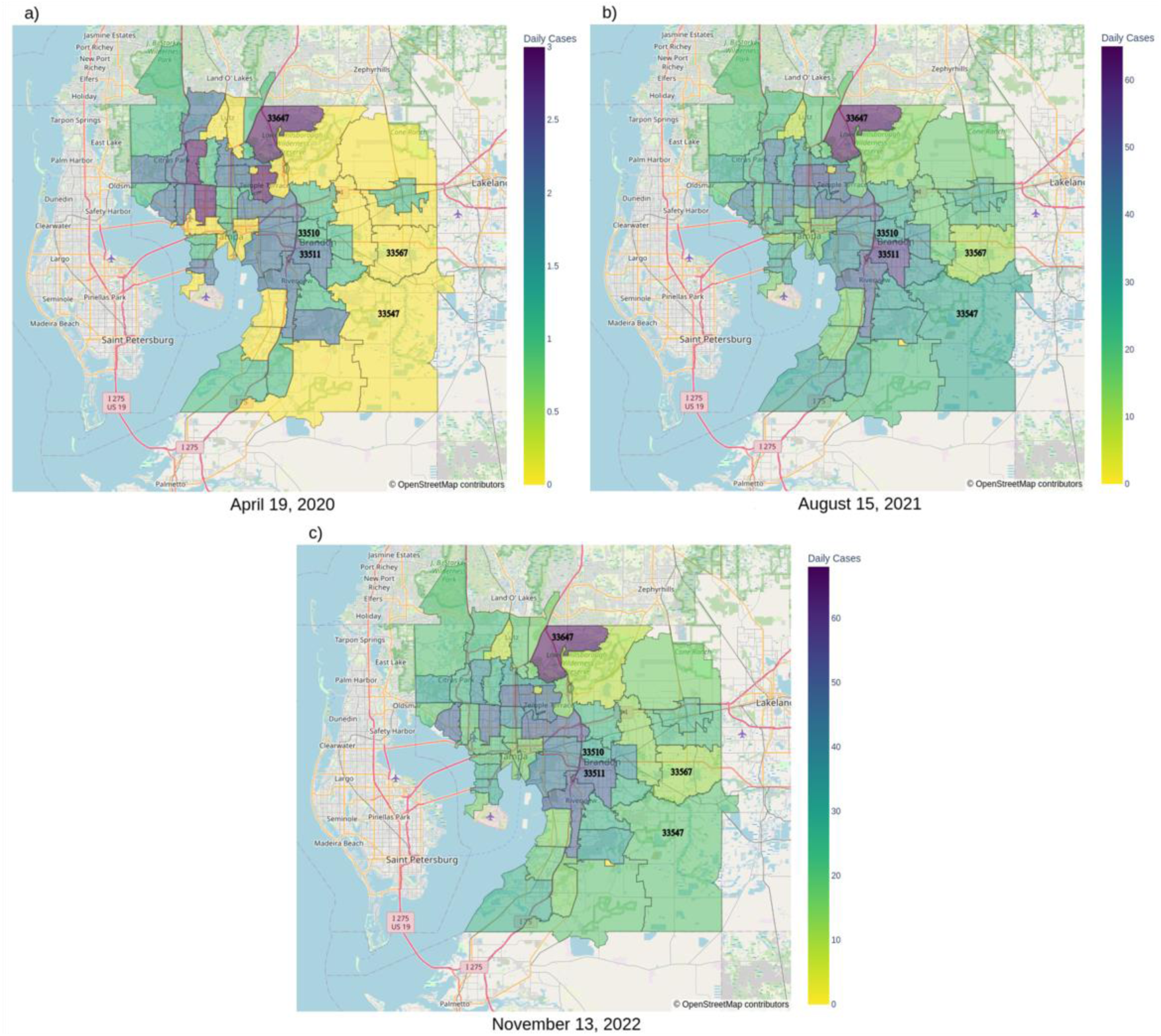
Maps of infection counts across zip codes on three dates (19^th^ April 2020, 15^th^ August 2021, and 13^th^ November 2022), showing spatial asynchrony between zip codes.

To further characterize the spatial synchronization in daily cases, we assessed the pattern of temporal correlations between the daily reported cases predicted for each pair of the 5 representative zip codes. We applied a moving 60-day time-window to perform the paired correlations to study the dynamics or evolution of zip code-level spatial synchronization in the cases [6]. This time-window length was arrived at by experimenting with different choices, which found that a 60-day window best helped in unveiling the synchronization patterns shown in Fig. 18. The results from this temporal correlation analysis highlighted the evolution of distinct synchronization patterns across the pairs of zip codes investigated. Notably, during the initial and Alpha variant waves, a low degree of synchronization was observed. This degree of synchronization gradually increased over time until the highest degree of synchronization was detected during the formation of the Delta and Omicron variant waves. The period post-Omicron, however, showed a return to lower levels of synchronization between the zip code pairs (Fig. 18). As noted above, this temporally-changing pattern of asynchrony and synchrony at different phases of the pandemic was also observed in the spatial distribution of evolving zip code-level cases in the county (Fig. 17). Several factors might interact to govern the spatial asynchrony-synchrony patterns observed in the results; during the early phases when most of the population is susceptible, variations in spatial case distribution as a result of non-uniform seeding of infectious cases, zip code-level population size, vulnerability to infection, and patterns of movements will govern the observed asynchronous pattern in cases. As waves develop, more and more susceptible individuals or agents come in contact with growing numbers of infecteds within and across zip codes to cause synchronization in the cases. During the post-omicron, synchrony is broken by the emergence of large but variable numbers of immunes in different zip codes (as a result of variations in the levels of infection experienced) forming barriers to transmission from lower numbers of infecteds to the remaining susceptibles.

**Fig. 18.**
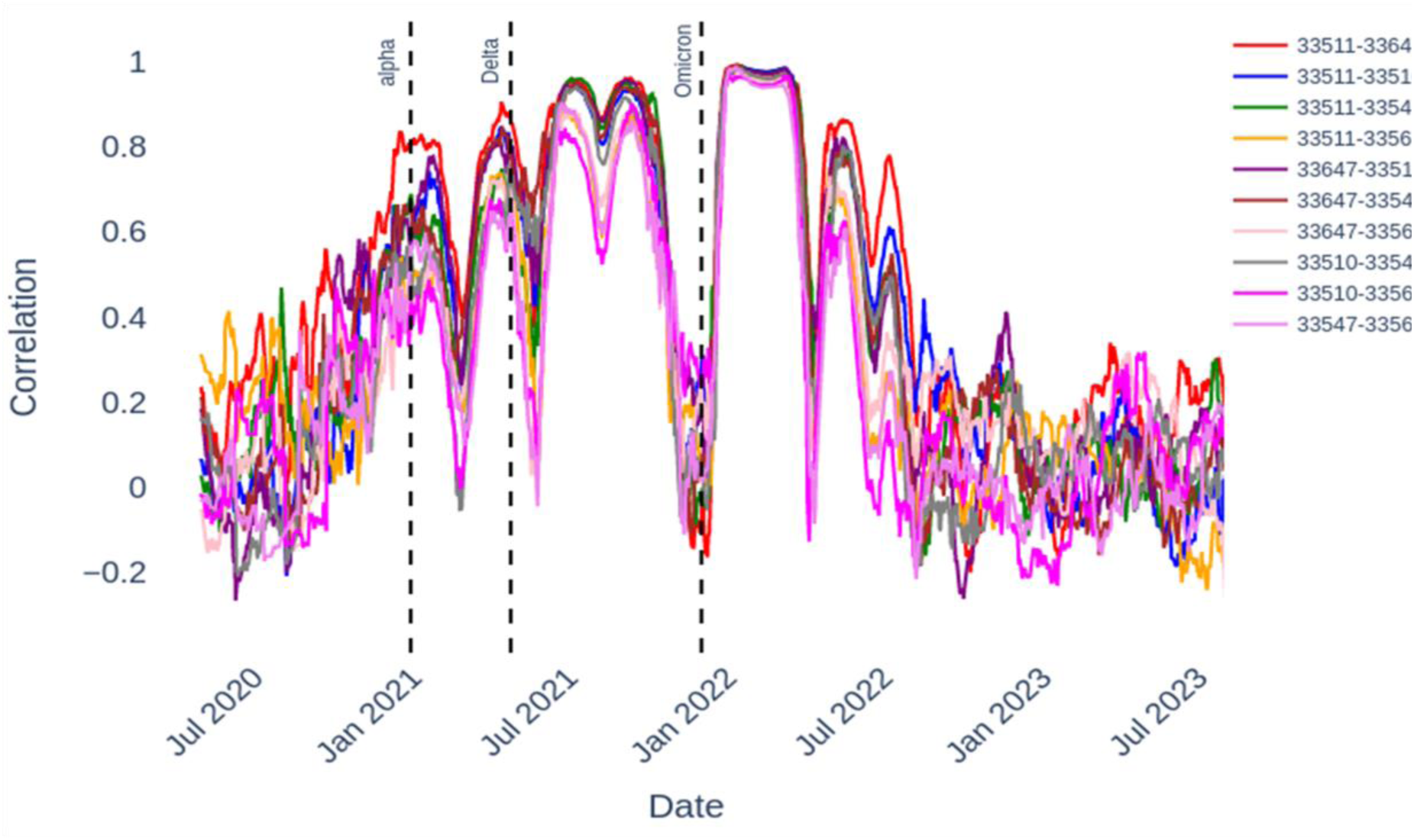
Moving window cross-correlations between the infection time series of the zip codes shown in the legend indicating emergent synchronization and asynchronization regions overtime.

Fig. 19 demonstrates another important utility of employing a place DT to run a disease ABM, viz. that it also allows investigation of the population attributes that may underlie the spatial (zip code level in the present case) distribution of disease transmission. The figure plots the fractions of agents exhibiting different attributes that may be associated with COVID-19 transmission in a sample of high incidence versus low incidence zip codes. The results show that among the agent attributes, the higher rate of transmission observed in the high incidence zip codes is associated with individuals belonging to >18 year groups, the Hispanic ethnic group, poorer income brackets and greater travel outside the resident zip code.

**Fig. 19.**
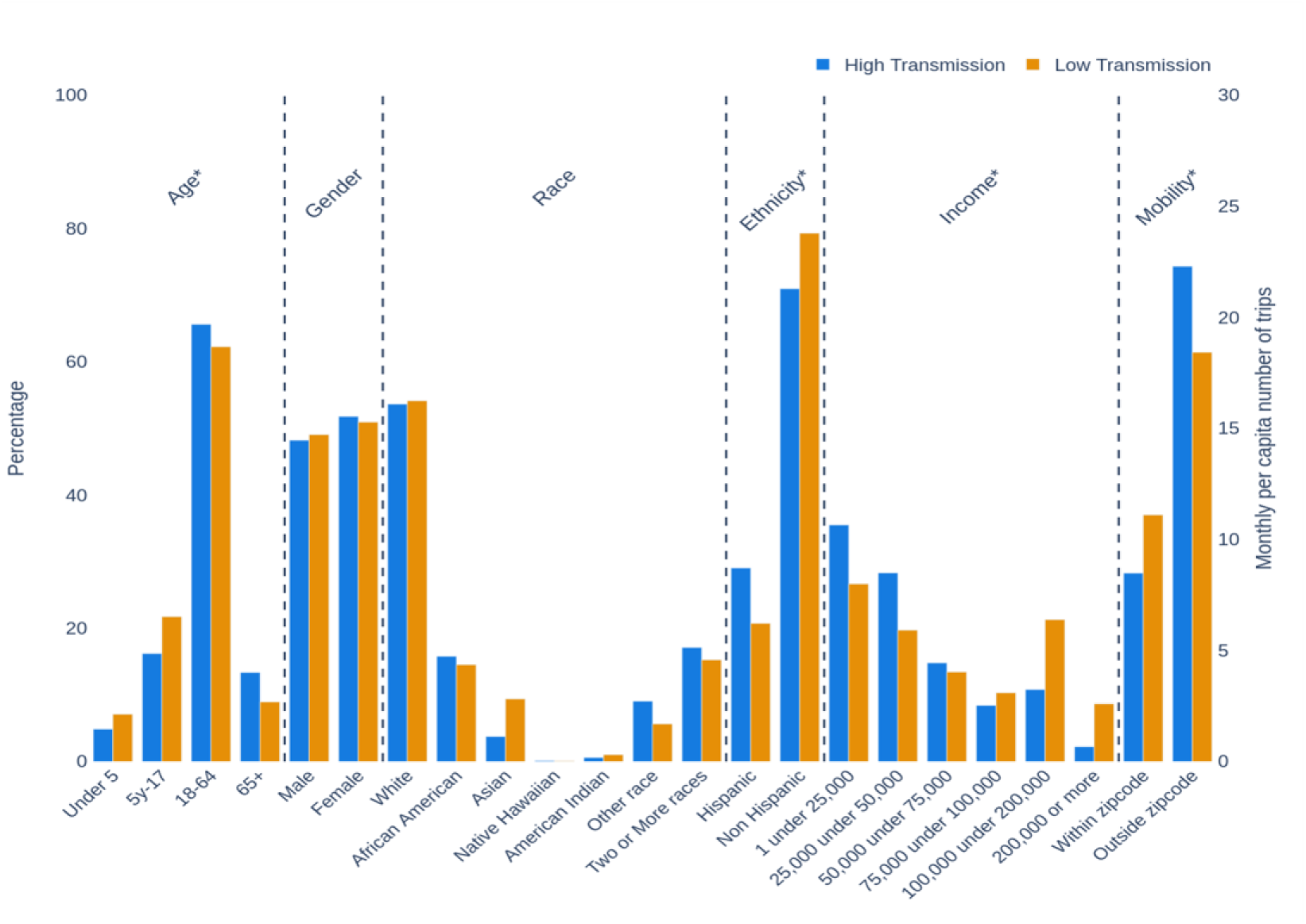
Population characteristics of low and high transmission zip codes.

### 3.3 Forecasts

We next used the model fitted to case data until end of July 2023 to generate near-future one-month ahead forecasts for the path of COVID-19 in the county. The results of the forecast shown in Fig. 20(a) indicates that viral transmission will assume a low-level oscillatory pattern in the near future. This implies that the infections will not fade away altogether despite the growth of high levels of naturally immune and vaccinated individuals over time (Fig. 20(b)). A further reason relates to the spatial evolution of immune agents across the county, which changes from an inhomogeneous pattern during the earlier stages of the pandemic to a high level more homogenous pattern by July 2023 (Fig. 21). This ensures that no local pockets of infections arise and is able to spread over time, thereby suppressing and maintaining infections at low levels globally. However, there is also a suggestion that due to waning of both natural and vaccine-induced immunity, combined with the immune escape potential of new variants as was reported for Omicron subvariants, there will be a continual risk of small flare-ups, as can be seen in Fig 20(a). An interesting feature of the results depicted in Fig 20(a) is that post-omicron, our model predictions are consistently higher than the reported case data despite fitting the omicron wave well. We suggest that this is mostly likely because case reporting began to become inconsistent during this stage raising important questions regarding whether the lower quality reported data or model predictions calibrated to past higher quality data offers a better guidance as to the later state of the pandemic.

**Fig. 20.**
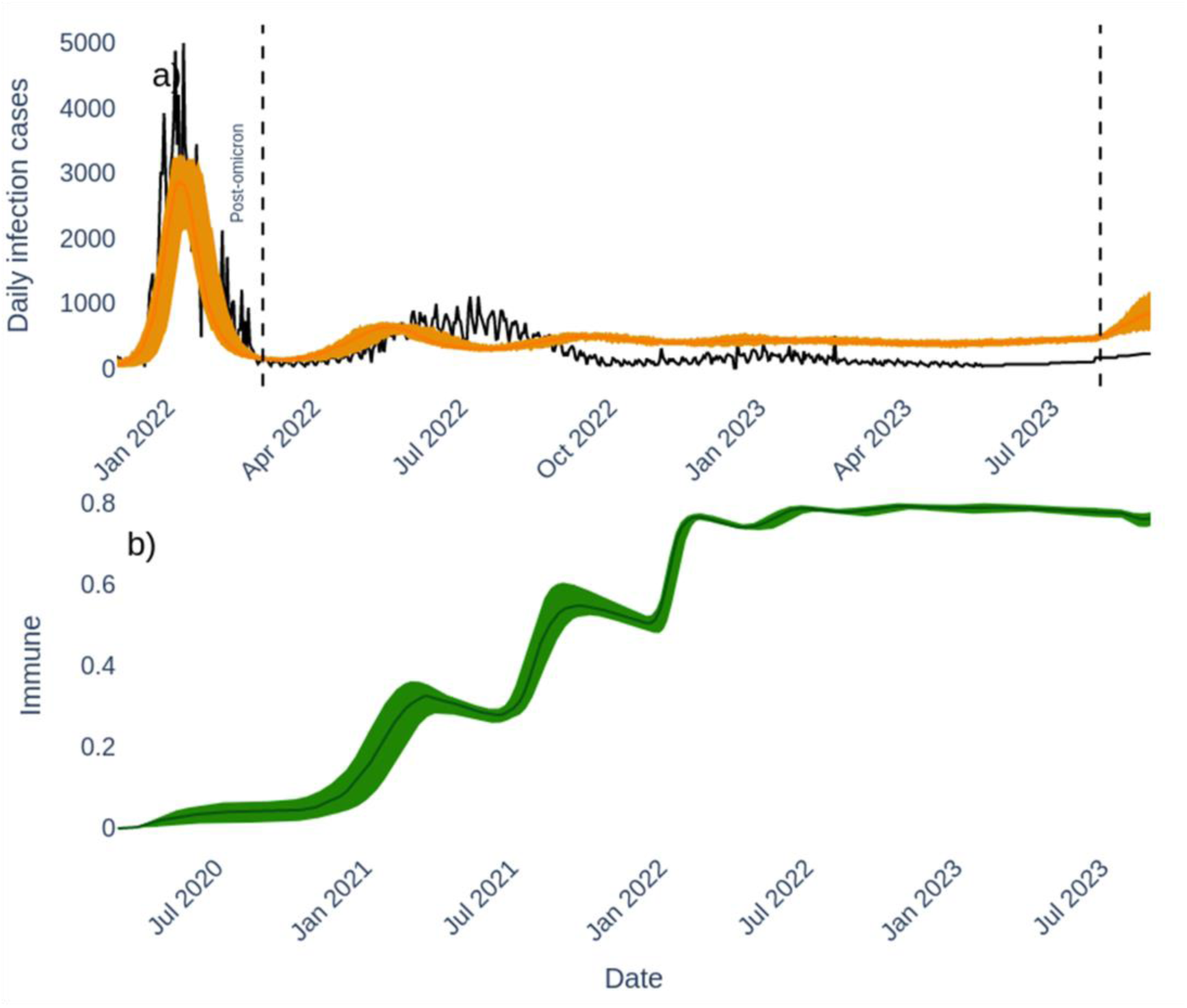
(a) Daily cases time series showing the time window from pre-omicron all the way into end of July 2023, and the period one month ahead to the end of August 2023. The dashed vertical lines indicate the beginning of the post-omicron and end of July 2023 periods respectively. (b) The daily change in the proportion of immune individuals over the course of the pandemic.

**Fig. 21.**
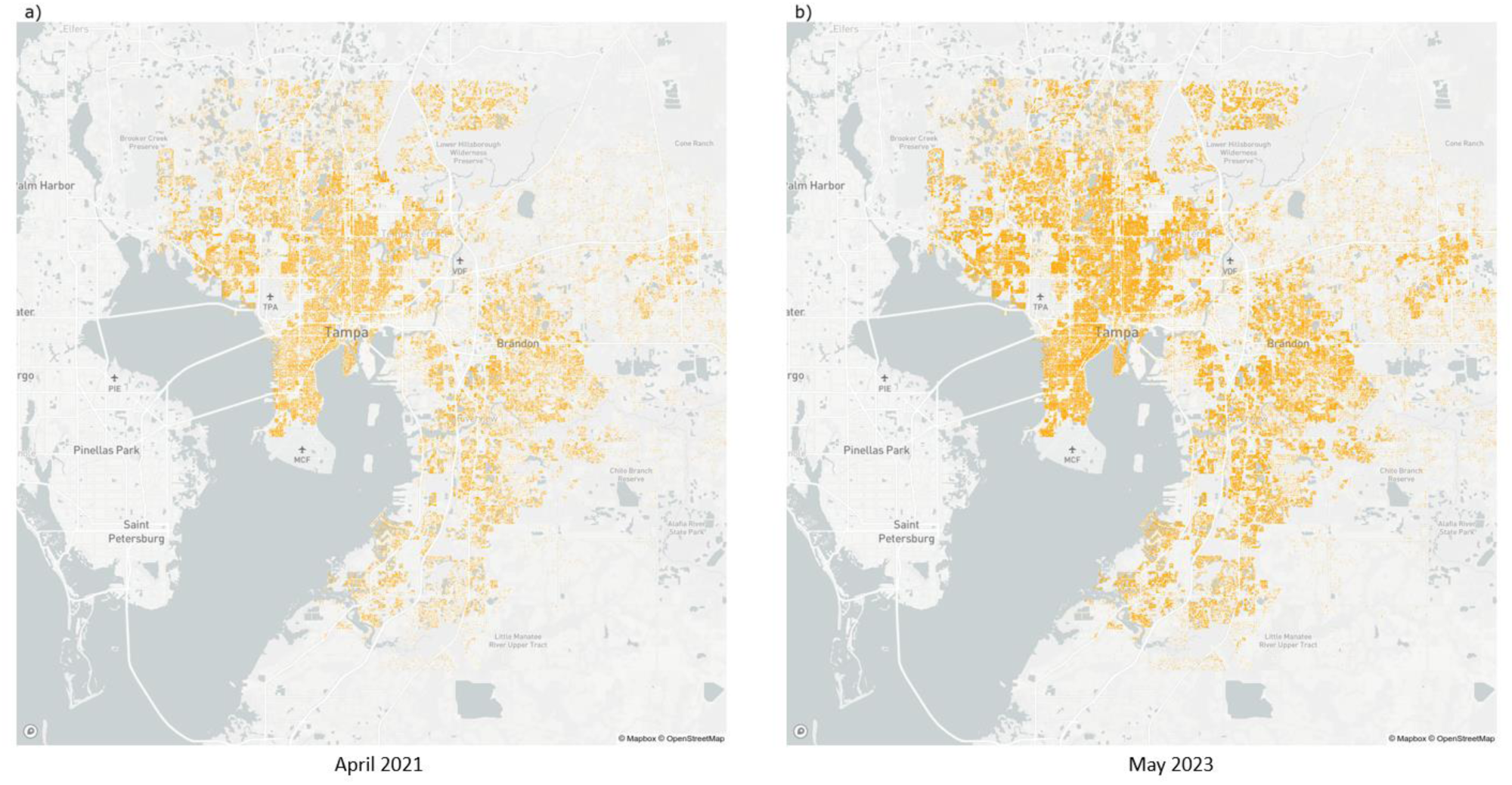
The modelled spatial distribution of immune agents in Hillsborough County in a) April 2021 and b) May 2023.

## 4 SUMMARY AND CONCLUSION

We describe how we have developed HPC-oriented workflows to implement a city-scale DT platform for the specification and execution of the key components of an ABM for simulating and controlling the spread of respiratory pandemics in complex urban settings, focusing here on the COVID-19 epidemic in Hillsborough County, Florida. The defining feature of the developed agent-based city DT for simulating the spread of such an epidemic is that it facilitates the construction of a fine-grained model of a city and its components in order to support more realistic data-driven disease modelling through the capture of the real-world factors (demographics, social behaviors and activities, locales of infections, and public interventions) that drive the rate and spatio-temporal spread of epidemics in populations. The workflows are also unique in that they facilitate the execution of large data-intensive steps that incorporate daily zip code and county-level surveillance and policy data, agent movements based on navigation/traffic data, dynamic agent behavioral responses to fluctuations in cases, individual agent susceptibility to the viral pathogen, and the arrival of viral variants differing in transmission and clinical characteristics. The workflows also additionally include post-simulation analytics for facilitating projections, and the making of sub-population and locational risk assessments. Computational efficiency and scalability constitute major issues when running large-scale ABM frameworks, and more so when these are embedded within a city DT. We have sought to resolve this issue using cloud-based HPC resources along with high parallelization of our code. Optimizing the trade-offs between performance and the computing cost of running HPC-supported digital-twin simulations on the cloud or on compute clusters is an ongoing challenge that needs to be resolved if we are to replace batch scheduling with on-demand requests of simulation runs [42, 49]; this will make it possible to eventually bring simulations closer to real-time. More realistic modelling of actual movements across a city by residents can also take advantage of new advances made in the use of real-time data from IoT sensors that integrates high-resolution infrastructure data and road user behaviors and vehicle movements across a city at various timepoints during the day [38]. This continuous synchronization of data will enable DT models to be continuously updated based on real-time conditions, making it an effective tool to run virtual simulations and scenario planning. Such systems will further increase computational complexity but will lead to a better bi-directional mapping of the real and digital worlds, the achievement of which will support better and smarter planning and management of health crises going forward.

This work arose in response to requests from county agencies to support their work on COVID-19 monitoring, risk assessment, and planning, and using the described workflows, we were able to provide uninterrupted bi-weekly simulations to guide their efforts for over a year from late 2021 to 2023. This experience and the results described here demonstrate that timely data-driven high resolution epidemic simulations using a coupled place DT-ABM platform is possible, and if the data inputs we have defined in the paper and increased compute power is made available, it is also scalable to larger regions. Indeed, we have already successfully applied the described data pipeline and compute workflows to the city of Miami (to be described in a following work), which amply validates this possibility. Finally, we note that while we have focused on respiratory epidemics in the present work and have validated our approach in the specific context of COVID-19 transmission, and associated intervention scenarios, our data pipeline and workflows are also designed on the principles of computational and model agility in such a way that the framework can be easily repurposed for other objectives and diseases [15]. We are exploring such extensions, for example, for simulating vector-borne diseases in Florida, and also as a simulation tool for aiding decision-making and preparedness for future epidemics in different national and global settings.

## Data Availability

All data produced in the present study are available upon reasonable request to the authors

