## Supplement document for "CitySEIRCast: An Agent-Based City Digital Twin for Pandemic Analysis and Simulation"

Complex & Intelligent Systems

Shakir Bilal[1], Wajdi Zaatour[1], Yilian Alonso Otano[1], Arindam Saha[1], Jun Kim[1], Kenneth Newcomb[1], Kim Soo[1], Raveena Ginjala[1,2], Derek Groen[3] and Edwin Michael*[1]

[1] Center for Global Health Infectious Disease Research, College of Public Health, University of South Florida, Tampa, FL 33612

[2]Department of Computer Science & Engineering, University of South Florida, Tampa, Florida 33612

[3] Modeling & Simulation Group, Department of Computer Science Brunel University London, Uxbridge UB8 3PH, United Kingdom

### Supplementary Information (SI)

This supplementary document contains the following details:

1. Table for age-dependent hospitalization, ICU and deaths.
2. Hillsborough lockdown and closures mandates for COVID-19.
3. Vaccine doses data and fitted polynomial curves.
4. Model complexity diagram based on data availability.

1. **Age-dependent risks**

The agent-based model uses age dependent hospitalization, ICU and death risks based on Table S1. Symptomatic individuals of different age groups either recover or get hospitalized, require ICU or die through different rates reflecting their immunity or comorbidity status.

Table S1. Age-dependent hospitalization, ICU and death parameters referenced from Enns et al. [1].

| Age group (years) | Percetange of confirmed cases that needed hospitalization | Percentage of hospitalized cases requiring ICU | ICU mortality rate (per 10 person-days) |
|---|---|---|---|
| 0-9 | 0.1% | 5.0% | 0.000 |
| 10-29 | 0.3% | 5.0% | 0.002 |
| 20-29 | 1.2% | 5.0% | 0.001 |
| 30-39 | 3.2% | 5.0% | 0.002 |
| 40-49 | 4.9% | 6.3% | 0.003 |
| 50-59 | 10.2% | 12.2% | 0.009 |
| 60-69 | 16.6% | 27.4% | 0.024 |
| 70-79 | 24.3% | 43.2% | 0.056 |
| 80> | 27.3% | 70.9% | 0.111 |

2. **Lockdown schedules in Hillsborough County, FL.**

The closure and lockdown schedules followed in Hillsborough County were collected from the mandates issued by the governors [2-6]. The detailed lockdown schedules that were applicable to places ranging from schools and workplaces to community places are shown in Figure S1. Since they include finer details, we distilled this information to reflect the closures vis-à-vis schools, workplaces, community place to inform our DT-ABM system. The distilled information that is used in the DT-ABM-system is shown in Table S3.

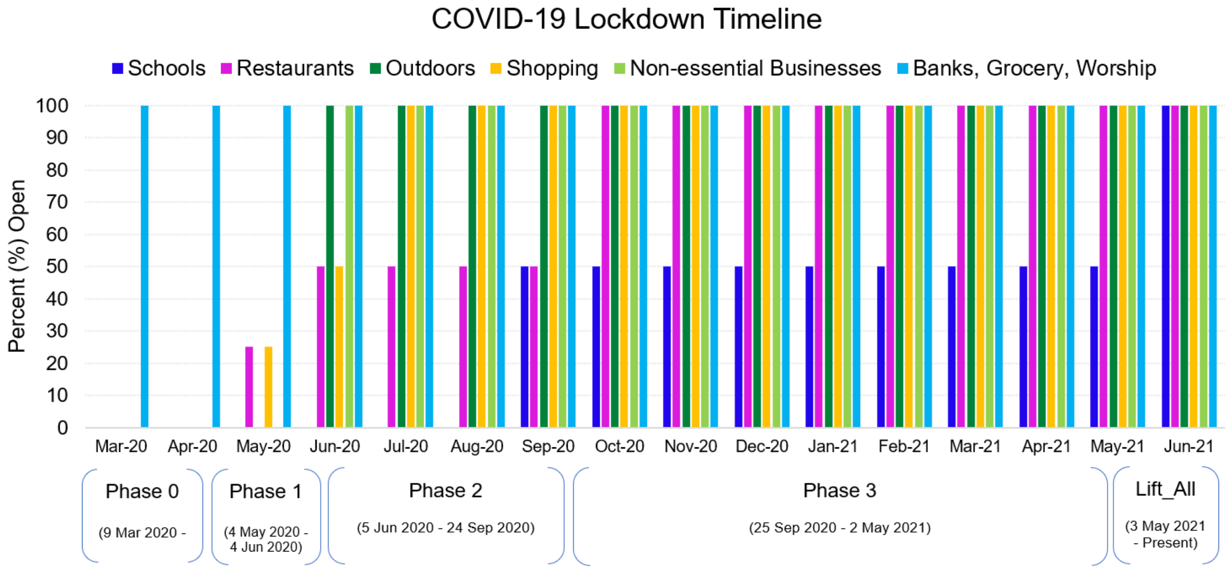

Figure S1. Lockdown timelines in Hillsborough County, Florida, during COVID-19 pandemic.

Table S3. Summary of lockdown schedule as imposed in Hillsborough County.

| Date Range | Office/Workplaces | Schools | Community |
|---|---|---|---|
| March 2020 - April 2020 | Open if essential (~10%) | Closed | Closed |
| April 2020 - May 2020 | 25% Open | Closed | 25% Open |
| May 2020 - July 2020 | 50% Open | Closed | 50% Open |
| August 2020 - April 202 | Fully Open | 50% Open | Fully Open |

| | |
|---|---|
| May 2021 – Present | All lockdown mandates are removed but the Google trends is used to mimic people adapting to changing Covid-19-related information. |

#### 3. Vaccine schedule and doses

We use the daily vaccine doses data available from CDC for Hillsborough County to implement vaccination in the simulator. Since there are gaps in the data we use polynomial fits to approximate the data and use the numbers suggested by these polynomials to vaccinate individuals in the agent-based model. The vaccine dose data and the fitted polynomial curves are shown in Figure S2. Furthermore, each dose type is divided into number of doses to different age groups as shown in Table S4.

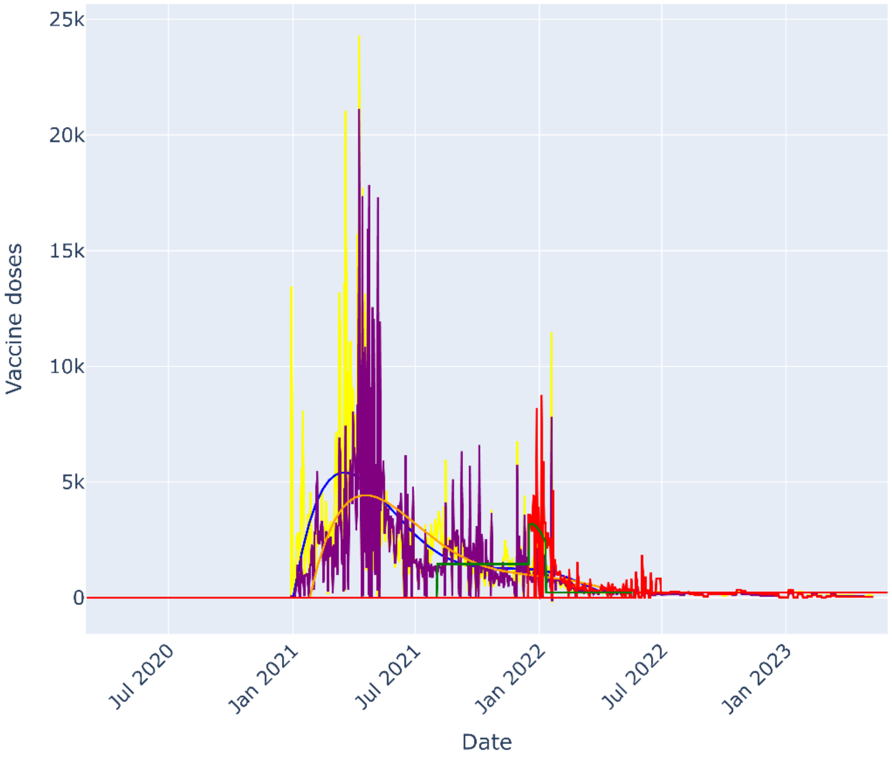

Figure S2. Daily vaccine numbers delivered in Hillsborough over time, starting with vaccine dose 1, dose 2, booster 1, booster 2.

Table S4.  The daily doses by age groups.

| Date range | Age groups (years) | Percent of Doses |
|---|---|---|
| January 21 - March 19, 2021 | 60+ | 100% |
| March 19 - April 5, 2021 | 30-60 | 34% |
| | 60+ | 66% |
| April 5 - December 31, 2021 | 10-30 | 33% |
| | 30-60 | 33% |
| | 60+ | 34% |
| January 1, 2022 - Present | <30 | 50% |
| | 30-60 | 25% |
| | 60+ | 25% |

4. **Model complexity**

The full DT-ABM relies on data availability to function. Based on the details of the available the complexity of the system increases. A summary of data availability and complexity is shown in Figure S3. As an example, if race/income data is available then individuals' race/income specific data is used in the DT-ABM. The output from DT-ABM analysis could then be used income and race-related risk factors.

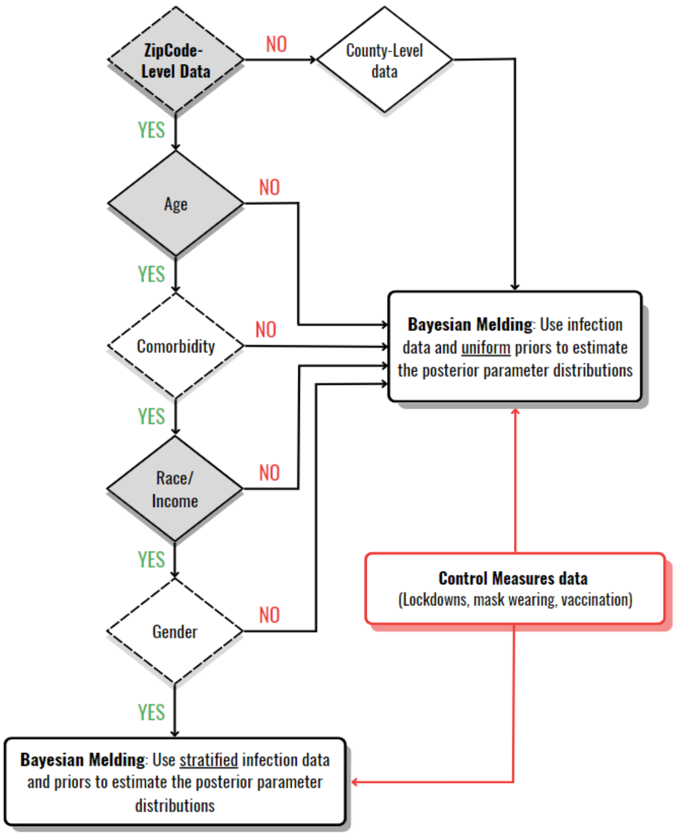

Figure S3: The complexity of the simulator increases as data for age-, comorbidity-, race- and gender-based transmission factors become available at the zip-code or county level.
